# Incorporating interactions into structured life course modelling approaches: A simulation study and applied example of the role of access to green space and socioeconomic position on cardiometabolic health

**DOI:** 10.1101/2023.01.24.23284935

**Authors:** Daniel Major-Smith, Tadeáš Dvořák, Ahmed Elhakeem, Deborah A. Lawlor, Kate Tilling, Andrew D. A. C. Smith

## Abstract

**Background:** Structured life course modelling approaches (SLCMA) have been developed to understand how exposures across the lifespan relate to later health, but have primarily been restricted to single exposures. As multiple exposures can jointly impact health, here we: i) demonstrate how to extend SLCMA to include exposure interactions; ii) conduct a simulation study investigating the performance of these methods; and iii) apply these methods to explore associations of access to green space, and its interaction with socioeconomic position, with child cardiometabolic health.

**Methods:** We used three methods, all based on lasso regression, to select the most plausible life course model: visual inspection, information criteria and cross-validation. The simulation study assessed the ability of these approaches to detect the correct interaction term, while varying parameters which may impact power (e.g., interaction magnitude, sample size, exposure collinearity). Methods were then applied to data from a UK birth cohort.

**Results:** There were trade-offs between false negatives and false positives in detecting the true interaction term for different model selection methods. Larger sample size, lower exposure collinearity, centering exposures, continuous outcomes and a larger interaction effect all increased power. In our applied example we found little-to-no association between access to green space, or its interaction with socioeconomic position, and child cardiometabolic outcomes.

**Conclusions:** Incorporating interactions between multiple exposures is an important extension to SLCMA. The choice of method depends on the researchers’ assessment of the risks of under-vs over-fitting. These results also provide guidance for improving power to detect interactions using these methods.

**Key messages:**

- In life course epidemiology, it is important to consider how multiple exposures over the lifespan may jointly influence health.
- We demonstrate how to extend current structured life course modelling approaches to include interactions between multiple different exposures.
- A simulation study comparing different methods to detect a true interaction effect found a trade-off between false positives and false negatives, suggesting that the optimal choice of method may depend on the researchers’ assessment of this trade-off (e.g., exploratory studies may prefer a greater risk of false positives, while confirmatory studies may prefer to minimise the risk of false positives).
- We identified key factors that improve power to detect a true interaction effect, namely larger sample sizes, centering exposures, lower exposure collinearity, continuous outcomes and larger interaction effect sizes.
- We applied these methods in a UK birth cohort (ALSPAC; Avon Longitudinal Study of Parents and Children), finding little-to-no evidence of an association between access to green space and its interaction with socioeconomic position on child BMI, obesity or blood pressure.

## Introduction

Life course epidemiology studies the effects on health of biological, social and environmental exposures during gestation, infancy, adolescence, adulthood, and across generations (Ben-Shlomo & Kuh 2002; Kuh *et al*. 2003). Over the last decade several structured life course modelling approaches (SLCMA; pronounced “slick-mah”) have been developed to help with the challenges of understanding these life course effects (Mishra *et al*. 2009; Smith *et al*. 2015, 2016, 2022; Howe *et al*. 2016; Madathil *et al*. 2018; Zhu *et al*. 2021). These approaches have largely focused on how to model the effect of one repeated exposure over the life course on an adult outcome, to distinguish for example between critical or sensitive periods, or cumulative effects. Little attention has been applied to two or more exposures and how these may jointly affect an outcome, despite the definition of life course epidemiology clearly highlighting the importance of multiple exposures jointly influencing health.

Here we extend these existing models by demonstrating how to incorporate interactions between multiple exposures in a SLCMA, allowing an exploration of how multiple exposures are associated with an outcome. This paper has three aims: i) describe how to extend existing structured life course models to include interaction terms between multiple exposures; ii) conduct a simulation study exploring how well this approach performs under a range of conditions; and iii) apply this approach in a UK birth cohort.

### Motivating Example

Throughout this paper we will use access to green space during pregnancy, infancy and early childhood, and its interaction with family socioeconomic position (SEP), and how these impact later child cardiometabolic outcomes (body mass index [BMI] and blood pressure), as a motivating example. BMI and blood pressure are key risk factors for cardiovascular disease progression (Berenson *et al*. 1998) which often manifest in childhood (Chen & Wang 2008; Singh *et al*. 2008). Understanding the risk factors – and potential interventions – for childhood obesity and hypertension is therefore a major public health concern (World Health Organization 2016). One potential modifiable risk factor is access to green space. Numerous studies have reported associations between access to green space and improved BMI and blood pressure in adults (James *et al*. 2015; Luo *et al*. 2020), although associations in children have been mixed (Bell *et al*. 2008; Wolch *et al*. 2011; Lovasi *et al*. 2013; Markevych *et al*. 2014; Picavet *et al*. 2016; Schalkwijk *et al*. 2018; Benjamin-Neelon *et al*. 2019; Bloemsma *et al*. 2019; Abbasi *et al*. 2020; Xiao *et al*. 2020; Warembourg *et al*. 2021; Jia *et al*. 2021; Cadman *et al*. 2022; Dzhambov *et al*. 2022; Luo *et al*. 2022). Interactions between access to green space and SEP on cardiovascular outcomes have received less attention, although some effect modification has been reported in both adults (James *et al*. 2015) and children (Schalkwijk *et al*. 2018). Previous studies have examined green space exposure at one time point, but, to the best of our knowledge, none have adopted a life course approach to identify how the association between green space and child BMI and blood pressure changes over infancy and childhood, or its interaction with SEP.

### The Structured Life Course Modelling Approach (SLCMA)

The first step in a SLCMA is to specify and encode potential life course hypotheses, which may either be ‘simple’ (encoded into one variable) or ‘compound’ (a combination of two or more encoded variables; (Mishra *et al*. 2009; Smith *et al*. 2015, 2016, 2022)). The selection of hypotheses could be exploratory or confirmatory; here we focus on exploratory analyses. Table 1 describes some common life course hypotheses and their extension to include interactions, and how they can be encoded where the exposures and confounders/covariates are binary variables. We use the term ‘confounder/covariate’ throughout, as the interaction term could include either the exposure and a confounder, or the exposure and an effect-modifier that is not a confounder. Similar encoding applies when the variables are continuous (Smith *et al*. 2016), but we introduce the key concepts here using binary variables.

**Table 1:**
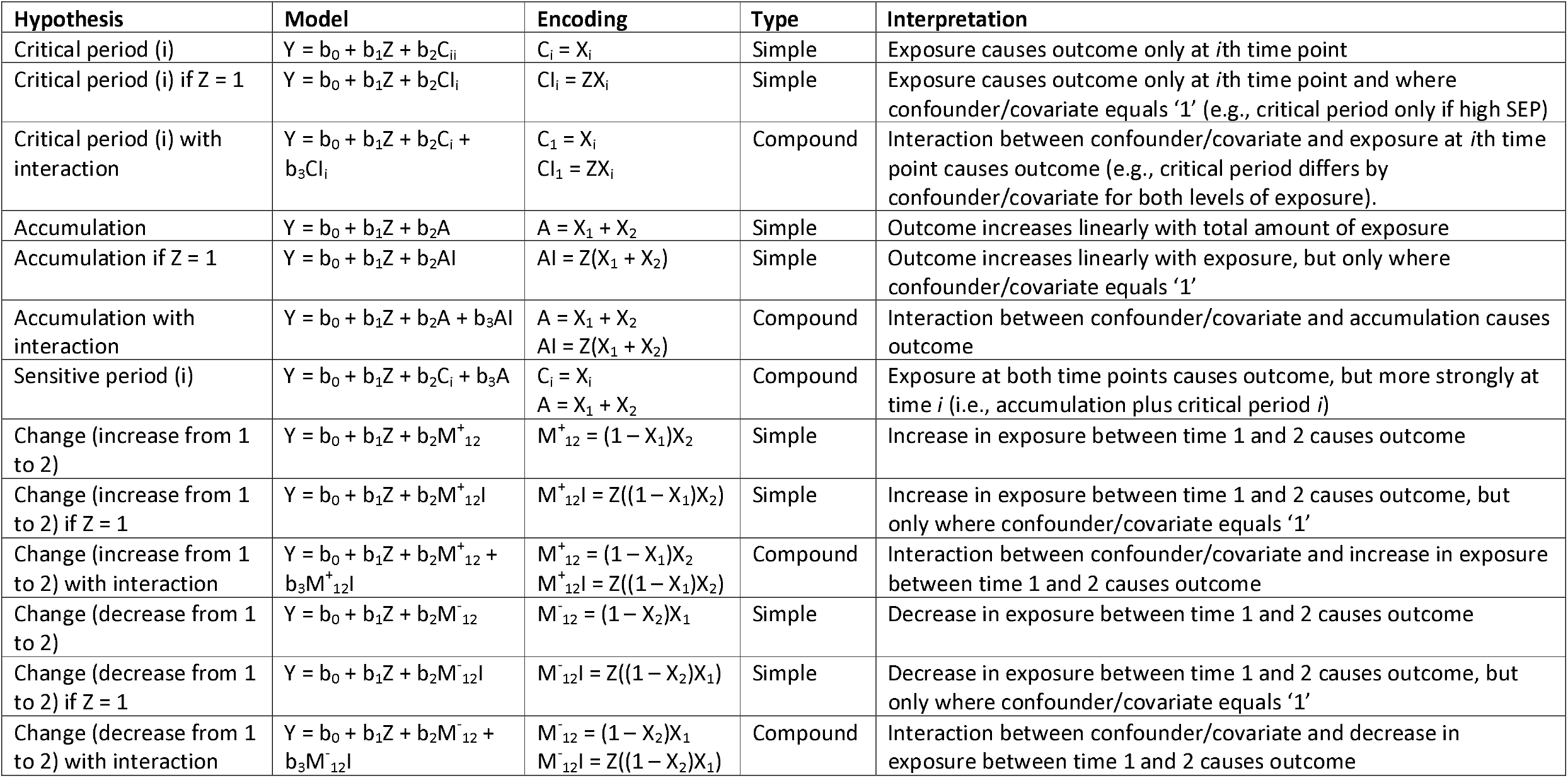
Examples of encoding hypotheses for structured life course models where exposure variables are binary and there are two exposure time-points. *X*_*i*_ is an indicator for the exposure at time-point *i*; *Z* indicates the confounder/covariate to be included in the interaction term. Note that as we have decided here that the confounder/covariate *Z* will be included in all models by default, it appears in all formulae in the ‘model’ column below. Not all possible life course model permutations are displayed here.

Once these hypotheses have been encoded, these terms are entered into a lasso regression model. Lasso models apply a penalty term (lambda) which constrains terms in the model to zero, an approach known as L1-regularisation. The initial lambda value is set so that there are no variables in the model; lambda is then decreased, allowing variables which explain the highest amount of the variation in the outcome into the model in a cumulative manner (Hastie *et al*. 2015). Previous structured life course approaches (Smith *et al*. 2015, 2016) have used an algorithm for this approach known as Least Angle Regression (LARS; (Efron *et al*. 2004)) which finds the best lasso model as lambda decreases. The first model contains the variable with the strongest association with the outcome, with later models containing additional variables added in a stepwise procedure. The choice of hypotheses is then based on an inspection of an ‘elbow plot’ (the proportion of variation explained after new variables are added), a formal lasso covariance hypothesis test as to whether inclusion of another variable improves model fit (Lockhart *et al*. 2014), or both. However, concerns have been raised about this lasso covariance test, especially regarding binary outcomes, and this approach is no longer recommended (Zhu *et al*. 2021).

Hence, there is currently no consensus on the optimal method for selecting the best fitting lasso model; we explore three possible approaches below, all of which are based on a standard lasso, rather than the LARS approach. These are:

1. Visual inspection. Examining the order in which encoded variables are entered into the model and the variance explained associated with each variable (similar to a LARS elbow plot). While this is a relatively straightforward approach, it is subjective, especially if the choice of best variable(s) is not clear.
2. Using a ‘relaxed lasso’ (Hastie *et al*. 2020) approach and comparing model fit using information criteria. After running the lasso model, take all model combinations found by the lasso, run a standard regression model on each (e.g., linear models for continuous outcomes, logistic regression for binary outcomes), and select the best-fitting model based on an information criterion which has a penalty for over-fitting (e.g., AIC [Akaike Information Criterion] or BIC [Bayesian Information Criterion]). Although both assess model fit, the AIC and BIC are calculated differently (Kuha 2004), with the BIC penalising complexity more than the AIC. We will use both the AIC and BIC here, and compare their performance.
3. Using a cross-validated lasso approach. Unlike an ordinary lasso which continuously improves model fit as lambda reaches 0, at each lambda value cross-validated lasso splits the data into *k* equal portions (here, *k* = 10) and calculates the mean prediction error. The lambda value which minimises the prediction error is deemed the ‘optimal’ model for predicting the outcome. We will compare two cross-validated methods for selecting the best-fitting model, one based on the model with the lowest mean prediction error and another selecting the model within 1 standard error of the minimum mean prediction error (which selects a sparser model and may avoid over-fitting; ((Hastie *et al*. 2015), pages 13-14)).

Note that the primary aim of this SLCMA as used in the simulation sections of this paper is not in estimation of the coefficients, but rather model selection and understanding the overall impact of the exposure on the outcome throughout the life course. For post-selection inference methods to calculate unbiased effect estimates, confidence intervals and *p*-values when using SLCMA – known as ‘selective inference’ – see; (Tibshirani *et al*. 2016; Zhu *et al*. 2021; Smith *et al*. 2022). Throughout this paper we focus on lasso approaches using the ‘glmnet’ package in R (Friedman *et al*. 2010).

### Worked Example

We first demonstrate these methods with a simple simulated dataset based on our motivating example, using BMI as our cardiometabolic outcome (see section S1 of the supplementary information for simulation details; simulated *n* = 10,000; all code is openly-available on GitHub, see the Data Availability section). In this simulation, there is one binary SEP covariate (1 = high SEP), three binary green space exposures (in pregnancy, at age 4, and at age 7; 1 = access to green space), and a continuous BMI outcome. We simulate that SEP causes all three green space exposures (higher SEP = greater access to green space) and the outcome (higher SEP = lower BMI), and is therefore a confounder of the green space-BMI relationship. In this example, we simulate a critical period where most recent access to green space at age 7 has a causal effect on BMI, but only when interacting with SEP, such that lower SEP and access to green space causes a reduction in BMI, but access to green space for those with higher SEP has no impact on BMI. This example is purely illustrative, although these effects may be plausible (James *et al*. 2015; Schalkwijk *et al*. 2018). The life course hypotheses encoded are described in table 2.

**Table 2:** Hypotheses encoded in our simulated worked example dataset. The variables in this dataset are: green1 (access to green space at time1/in pregnancy), green2 (access to green space at time 2/age 4), green3 (access to green space at time 3/age 7) and high_sep (binary marker for high socioeconomic position). The confounder ‘high_sep’ is included in all models. ‘Simple’ models can be encoded in just one additional variable, while ‘compound’ hypotheses combine two or more additional encoding variables. In this simulated example, the compound hypothesis ‘crit3 + int3’ (critical period at time 3/age 7 with an interaction with SEP) is the true model.

| Hypothesis | Encoding | Type |
| --- | --- | --- |
| Critical period (1) | crit1 = green1 | Simple |
| Critical period (1) only if high SEP | int1 = high_sep * green1 | Simple |
| Critical period (1) with SEP interaction | crit1 + int1 | Compound |
| Critical period (2) | crit2 = green2 | Simple |
| Critical period (2) only if high SEP | int2 = high_sep * green2 | Simple |
| Critical period (2) with SEP interaction | crit2 + int2 | Compound |
| Critical period (3) | crit3 = green3 | Simple |
| Critical period (3) only if high SEP | int3 = high_sep * green3 | Simple |
| Critical period (3) with SEP interaction | crit3 + int3 | Compound |
| Accumulation | accumulation = green1 + green2 + green3 | Simple |
| Accumulation only if high SEP | int_accum = high_sep * accumulation | Simple |
| Accumulation with SEP interaction | accumulation + int_accum | Compound |
| Sensitive period (1) | crit1 + accumulation | Compound |
| Sensitive period (2) | crit2 + accumulation | Compound |
| Sensitive period (3) | crit3 + accumulation | Compound |
| Change (more green space from 1 to 2) | green_inc12 = (1 – green1) * green2 | Simple |
| Change (more green space from 1 to 2) only if high SEP | green_inc12_int = high_sep * green_inc12 | Simple |
| Change (more green space from 1 to 2) with SEP interaction | green_inc12 + green_inc12_int | Compound |
| Change (less green space from 1 to 2) | green_dec12 = (1 – green2) * green1 | Simple |
| Change (less green space from 1 to 2) only if high SEP | green_dec12_int = high_sep * green_dec12 | Simple |
| Change (less green space from 1 to 2) with SEP interaction | green_dec12 + green_dec12_int | Compound |
| Change (more green space from 2 to 3) | green_inc23 = (1 – green2) * green3 | Simple |
| Change (more green space from 2 to 3) only if high SEP | green_inc23_int = high_sep * green_inc23 | Simple |
| Change (more green space from 2 to 3) with SEP interaction | green_inc23 + green_inc23_int | Compound |
| Change (less green space from 2 to 3) | green_dec23 = (1 – green3) * green2 | Simple |
| Change (less green space from 2 to 3) only if high SEP | green_dec23_int = high_sep * green_dec23 | Simple |
| Change (less green space from 2 to 3) with SEP interaction | green_dec23 + green_dec23_int | Compound |

Figure 1 shows the results of this simulated lasso model. The order in which variables are entered is displayed from left to right, with a measure of model fit (deviance ratio) on the *y*-axis (for a detailed interpretation of this plot, see the figure 1 legend). The first variable entered in the model was ‘crit3’ (critical period at time 3/age 7), followed by ‘int3’ (interaction between SEP and critical period at time 3/age 7), and there appears to be an appreciable increase in model fit associated with both variables before additional variables are added (approx. 1.5% for both variables). As the line is largely horizontal after these two variables have been added, this suggests that the inclusion of additional variables does not noticeably improve model fit. This indicates that critical period at time 3/age 7 and its interaction with SEP have the strongest association with the outcome, and that all other variables have a much weaker/null association.

**Figure 1:**
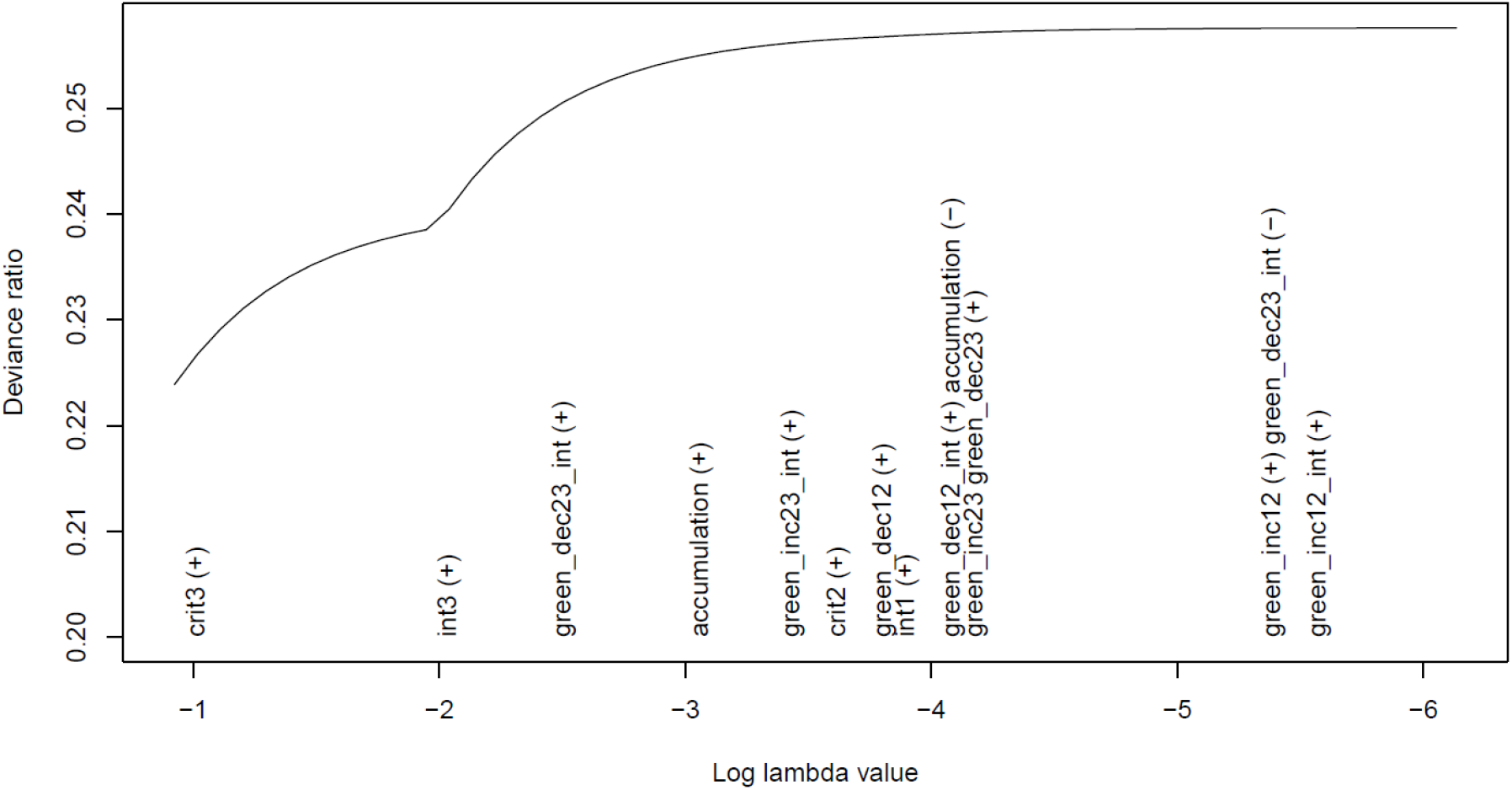
Results of the lasso from the simulated worked example. This plot displays the log lambda value on the *x*-axis (the lasso penalty term which allows more variables to enter the model) and the absolute deviance ratio of the lasso model associated with each lambda value on the *y*-axis (as a measure of model fit). As the variables are added to/removed from the model they appear on the *x*-axis, moving from left to right (with “(+)” indicating addition, and “ (.) ” indicating removal). The covariate/confounder ‘high_sep’ does not appear in this plot as it was constrained to be included in all models by default. This plot shows that the first encoded variable to enter the model was access to green space at critical period 3/age 7 (‘crit3’). This variable was then associated with an increase in the deviance ratio by approximately 1.5% until the next encoded variable was added, that of an interaction between critical period 3/age 7 and SEP (‘int3’). After this variable was added, there was another increase in the deviance ratio of approximately 1.5% when the next encoded variable was added, that of ‘green_dec23_int’ (the interaction term between a decrease in green space between times 2 and 3 and SEP). When this and subsquent variables were added, the improvements in the deviance ratio were minimal – as indicated by the largely horizontal line by this point – suggesting that none of these encoded variables were strongly associated with the outcome BMI (for an explanation of all other encoded variables in this plot, see table 2). These results suggest that the combination of ‘crit3’ and ‘int3’ are the best fit to the data, with all other variables having a much weaker/null association with the outcome, just as was simulated.

Next, we tested these patterns more formally using the relaxed lasso approach. The lowest AIC and BIC values were found for the true model, containing both critical period at time 3/age 7 and its interaction with SEP (in addition to the ‘high_sep’ confounder/covariate included in all models by default). As with the more subjective visual inspection method, we again reach the conclusion that the compound hypothesis of recent access to green space by SEP interaction is the best model.

Finally, we can use cross-validated lasso for model selection. The model within one standard error of the model with the lowest mean-squared error correctly identified critical period at time 3/age 7 and its interaction with SEP (in addition to ‘high_sep’) as the best-fitting model. The cross-validated model with the lowest mean prediction error included both of these variables, in addition to nine other encoded variables (figure S1), indicating overfitting. All three of these methods – except for cross-validated lasso using the model with the minimum mean prediction error – therefore provide broadly consistent results that match the simulated model.

### Simulation study

The previous section demonstrated how to extend SLCMA to include multiple exposures and interaction terms. However, the conditions under which these approaches identify the correct interaction term is unknown, especially given known issues of low statistical power for interactions (Brookes *et al*. 2004; Blake & Gangestad 2020). We conducted a formal simulation study (using the ADEMP approach; (Morris *et al*. 2019)) to explore how several factors impact the probability of the SLCMA methods detecting the true interaction term (i.e., the correct SEP-interaction, regardless of other terms in the final model).

#### Aim

Assess how often SLCMA identifies the true interaction term, while varying the following factors:

- Sample size: 1,000 vs 10,000
- Exposure variables: binary vs continuous (for binary exposures we used the life-course hypotheses encoded in table 2, while for continuous exposures we used the hypotheses encoded in table S1)
- Centering exposure variables: no vs yes (centering exposures may reduce the collinearity between main effects and interaction terms (Afshartous & Preston 2011; Iacobucci *et al*. 2016))
- Collinearity between exposure variables: low vs high (higher collinearity may make it more difficult for the model to distinguish between different life course hypotheses)
- Outcome variable: binary vs continuous (analyses with binary outcomes tend to have lower power (Altman & Royston 2006))
- Size of interaction term: none vs very small vs small vs moderate vs large vs very large (ranked on a relative scale)
- Life course hypothesis interaction: critical period at time 3 vs accumulation vs change from time 2 to time 3

#### Data-generating mechanism

We used the same data-generating mechanism as above to generate all exposures, with SEP as a binary variable. Simulating the outcome BMI depended on the life course hypothesis being assessed, i.e., an interaction between SEP and either critical period at time 3, accumulation or change from time 2 to 3. See supplementary information section S2 for additional detail on these simulation parameters and how they were selected.

#### Estimand

The target estimand was the inclusion of the correct SEP-interaction term being selected in the final model.

#### Methods

We used the relaxed lasso approach (using both AIC and BIC) and cross-validated lasso (using both minimum mean-squared error [MSE] and within 1 standard error of the MSE) for model selection.

#### Performance measure

The percentage of simulations (out of 1,000) that selected the true interaction term.

The results of this simulation study across all simulation parameters, for each SLCMA method, interaction strength, and life course interaction model, are displayed in table 3. As the strength of the interaction increased more models selected the correct interaction term. The relaxed lasso AIC and cross-validated minimum MSE methods were more likely to select the target interaction (even frequently in the ‘no interaction’ scenario), while the relaxed lasso BIC and cross-validated 1SE approaches were less likely to select the interaction, even with larger interaction coefficients (with 1SE cross-validation less likely to detect an interaction than relaxed BIC). The relaxed AIC and cross-validated minimum MSE methods are therefore more likely to detect a true interaction term, but at the expense at an increased risk of false positives, while the reverse is true for the relaxed BIC and cross-validated 1SE approaches.

**Table 3:**
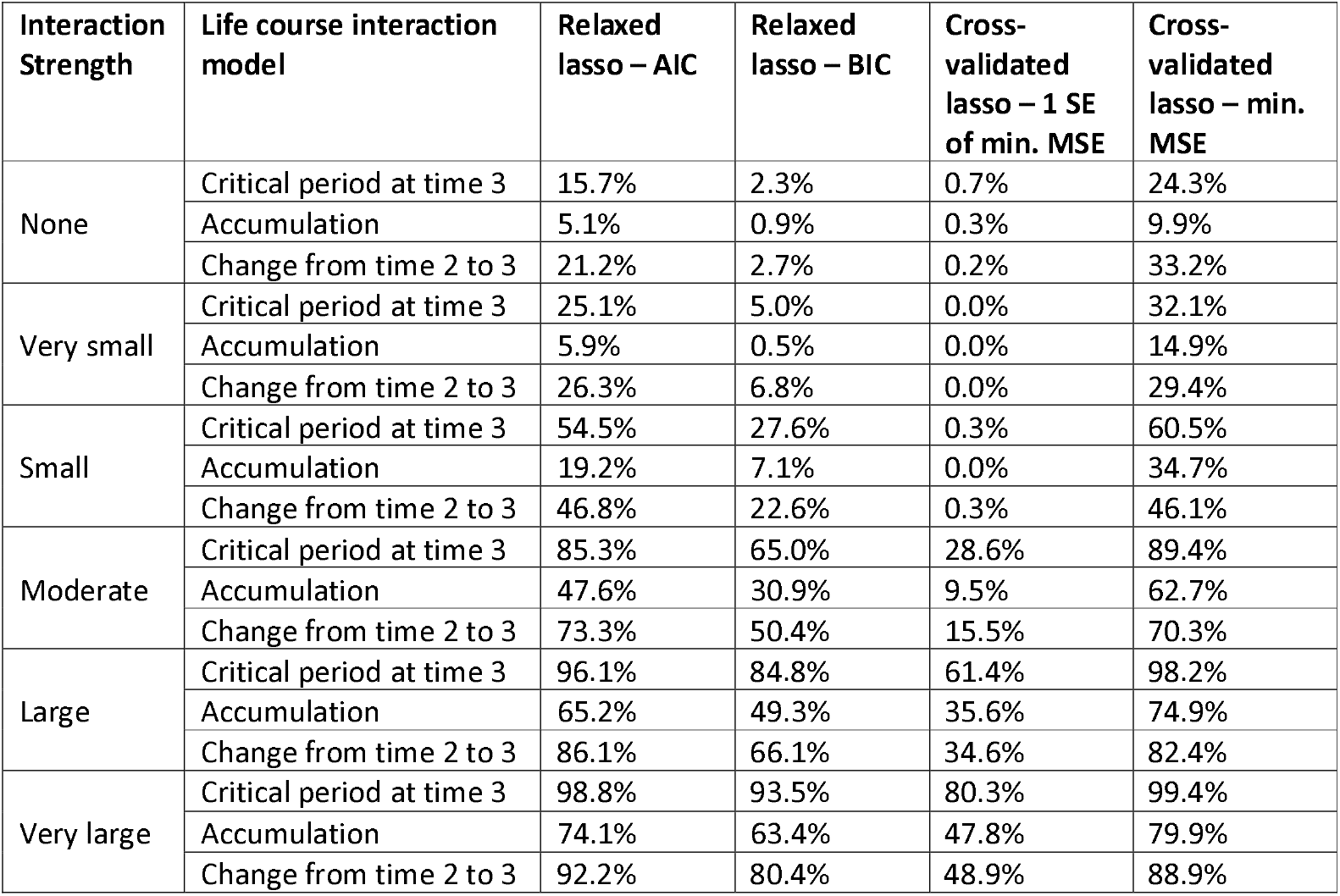
Summary of overall simulation results for each SLCMA method, each interaction strength, and each life course interaction model (*n* simulations = 32; *n* iterations per simulation = 1,000). Results show the percent of simulations in which the correct interaction term was selected. Note that in the rows for interaction strength of ‘None’, technically this is not the ‘correct’ interaction as no interaction was present (i.e., 15.7% of simulations using the AIC relaxed lasso method included the interaction term for the critical period at time 3 model, even though no interaction was simulated). For full details on the simulation parameters, see section S2 of the supplementary information. SE = Standard error; MSE = Mean-squared error.

| Interaction Strength | Life course interaction model | Relaxed lasso – AIC | Relaxed lasso – BIC | Cross-validated lasso – 1 SE of min. MSE | Cross-validated lasso – min. MSE |
| --- | --- | --- | --- | --- | --- |
| None | Critical period at time 3 | 15.7% | 2.3% | 0.7% | 24.3% |
|  | Accumulation | 5.1% | 0.9% | 0.3% | 9.9% |
|  | Change from time 2 to 3 | 21.2% | 2.7% | 0.2% | 33.2% |
| Very small | Critical period at time 3 | 25.1% | 5.0% | 0.0% | 32.1% |
|  | Accumulation | 5.9% | 0.5% | 0.0% | 14.9% |
|  | Change from time 2 to 3 | 26.3% | 6.8% | 0.0% | 29.4% |
| Small | Critical period at time 3 | 54.5% | 27.6% | 0.3% | 60.5% |
|  | Accumulation | 19.2% | 7.1% | 0.0% | 34.7% |
|  | Change from time 2 to 3 | 46.8% | 22.6% | 0.3% | 46.1% |
| Moderate | Critical period at time 3 | 85.3% | 65.0% | 28.6% | 89.4% |
|  | Accumulation | 47.6% | 30.9% | 9.5% | 62.7% |
|  | Change from time 2 to 3 | 73.3% | 50.4% | 15.5% | 70.3% |
| Large | Critical period at time 3 | 96.1% | 84.8% | 61.4% | 98.2% |
|  | Accumulation | 65.2% | 49.3% | 35.6% | 74.9% |
|  | Change from time 2 to 3 | 86.1% | 66.1% | 34.6% | 82.4% |
| Very large | Critical period at time 3 | 98.8% | 93.5% | 80.3% | 99.4% |
|  | Accumulation | 74.1% | 63.4% | 47.8% | 79.9% |
|  | Change from time 2 to 3 | 92.2% | 80.4% | 48.9% | 88.9% |

Variation between the life course interaction models was also apparent, with critical period interactions more likely to be selected than change interactions, and accumulation interactions least likely to be selected. This is likely due to the accumulation variables having high collinearity with the critical period variables, and therefore having less power to distinguish between these competing hypotheses.

This summary masks substantial variation, with methods performing well for some combinations of simulation parameters, and poorly for others. Focusing on the moderate interaction scenario, there is variation in the power to detect the correct interaction term (table 4). Overall, power was much greater with a larger sample size and continuous rather than binary outcomes. For other simulation parameters, differences were either smaller or varied by the life course interaction model. For instance, whether the exposures were binary or continuous made little difference for the critical period model, but power to detect an accumulation interaction was greater with binary exposures, while for the change interaction continuous exposures had greater power. High exposure collinearity greatly reduced the power of the accumulation and change models, while for the critical period models differences in collinearity were more modest. Centering the encoded variables increased power in the accumulation and critical period settings, but not in the change scenario.

**Table 4:** Summary of simulation results for each SLMCA method and each life course interaction models, under the ‘moderate interaction’ scenario, with results separated by each of the simulation parameters varied (*n* simulations = 32; *n* iterations per simulation = 1,000). Results show the percent of simulations in which the correct interaction term was selected (as there were 32 simulations, each row here contains the average over 16 simulations). For full details on the simulation parameters, see section S2 of the supplementary information. SE = Standard error; MSE = Mean-squared error.

| Simulation parameter | Life course interaction model | Relaxed lasso – AIC | Relaxed lasso – BIC | Cross-validated lasso – 1 SE of min. MSE | Cross-validated lasso – min. MSE |
| --- | --- | --- | --- | --- | --- |
| *Sample size* |  |  |  |  |  |
| 1,000 | Critical period at time 3 | 72.0% | 33.8% | 2.5% | 78.8% |
|  | Accumulation | 20.9% | 4.6% | 0.2% | 36.6% |
|  | Change from time 2 to 3 | 56.8% | 28.0% | 2.5% | 53.6% |
| 10,000 | Critical period at time 3 | 98.6% | 96.2% | 54.7% | 100.0% |
|  | Accumulation | 74.3% | 57.1% | 18.8% | 88.7% |
|  | Change from time 2 to 3 | 89.8% | 72.8% | 28.5% | 87.0% |
| *Exposure* |  |  |  |  |  |
| Binary | Critical period at time 3 | 86.4% | 69.6% | 34.7% | 89.1% |
|  | Accumulation | 56.7% | 39.5% | 16.8% | 66.4% |
|  | Change from time 2 to 3 | 61.5% | 35.2% | 10.3% | 56.3% |
| Continuous | Critical period at time 3 | 84.3% | 60.4% | 22.4% | 89.7% |
|  | Accumulation | 38.4% | 22.3% | 2.2% | 58.9% |
|  | Change from time 2 to 3 | 85.0% | 65.6% | 20.7% | 84.4% |
| *Collinearity of exposures* |  |  |  |  |  |
| Low | Critical period at time 3 | 90.4% | 69.5% | 30.0% | 90.9% |
|  | Accumulation | 60.5% | 42.4% | 6.6% | 69.4% |
|  | Change from time 2 to 3 | 91.0% | 75.7% | 30.9% | 89.8% |
| High | Critical period at time 3 | 80.2% | 60.5% | 27.1% | 87.9% |
|  | Accumulation | 34.7% | 19.4% | 12.4% | 55.9% |
|  | Change from time 2 to 3 | 55.6% | 25.2% | 0.1% | 50.9% |
| *Encoded variables centered* |  |  |  |  |  |
| No | Critical period at time 3 | 87.1% | 65.6% | 27.6% | 89.0% |
|  | Accumulation | 38.8% | 24.1% | 7.7% | 57.3% |
|  | Change from time 2 to 3 | 74.1% | 50.8% | 15.4% | 70.8% |
| Yes | Critical period at time 3 | 83.5% | 64.4% | 29.6% | 89.8% |
|  | Accumulation | 56.3% | 37.7% | 11.3% | 68.1% |
|  | Change from time 2 to 3 | 72.4% | 50.0% | 15.6% | 69.8% |
| *Outcome* |  |  |  |  |  |
| Continuous | Critical period at time 3 | 92.6% | 77.1% | 42.7% | 97.5% |
|  | Accumulation | 56.2% | 38.6% | 17.1% | 73.5% |
|  | Change from time 2 to 3 | 82.4% | 59.9% | 24.7% | 79.8% |
| Binary | Critical period at time 3 | 78.0% | 52.9% | 14.4% | 81.2% |
|  | Accumulation | 38.9% | 23.2% | 2.0% | 51.8% |
|  | Change from time 2 to 3 | 64.2% | 41.0% | 6.4% | 60.9% |

As it may not be possible to directly compare power between continuous and binary outcomes because the interaction terms are not equivalent, these results are repeated in tables S2 and S3, split by continuous vs binary outcome. Results are qualitatively similar to the combined results reported here.

### Applied example in a UK birth cohort

We used data from the Avon Longitudinal Study of Parents and Children (ALSPAC) as an illustrative example of how to apply the SLCMA methods detailed above. Our substantive research question concerns the impact of access to green space, and its potential interaction with SEP, on later child BMI (both continuous and a binary of measure of obesity) and blood pressure. We used ‘access to green space’ as our exposure, measured during pregnancy, age 4 and age 7 (this was a binary variable indicating whether there was a green space >5,000 m^2^ within 300 meters of their home). Our outcomes were BMI, overweight/obese, systolic blood pressure (SBP) and diastolic blood pressure (DBP) measured at age 7. We used parental education as our binary SEP interaction term (coded as ‘O-level or lower’ [lower SEP] vs ‘A-level or higher’ [higher SEP]). Maternal ethnicity, the sex of the child, and the age of child at the time of outcome measurement were included as confounders/covariates (see figure S2 for our hypothesised Directed Acyclic Graph displaying the assumed causal relations between variables). As all exposures were binary, hypotheses were encoded as described in table 2. Additional details of the study population, variable selection and analysis methods can be found in section S3 of the supplementary information.

At each of the time points approximately 75% of children had access to a green space (table S4), while variation in this exposure over time was relatively low, with 70% of children having access to a green space at all three time points and 17% having no access at all time points (table S5 and figure S3). Correlations between access to green space for adjacent time-points were high (*r* = 0.76 between pregnancy and age 4; *r* = 0.86 between age 4 and age 7). Descriptive statistics for the BMI, obesity and blood pressure outcomes, SEP confounders/interaction variables and the other confounders/covariates can be found in table S6.

We first present results where BMI was the outcome. Visual inspection indicated that inclusion of the first variables in the model – an interaction between SEP and critical period at age 4 (‘int2’) and an increase in access to green space between times 2 and 3 (‘green_inc23’) – had little association with the outcome as there was minimal improvement in model fit (figure 2). This was confirmed by both other methods, with the relaxed lasso (AIC and BIC) and cross-validated lasso (minimum MSE and 1SE of MSE), selecting the covariate-only model as the best fit to the data (table S7). Given the rather limited variation in this green space exposure over time, we also explored whether using just two time-points (pregnancy and age 7) would give different results; results were qualitatively similar (figure S4), suggesting that results from the models using all three time-points are robust.

**Figure 2:**
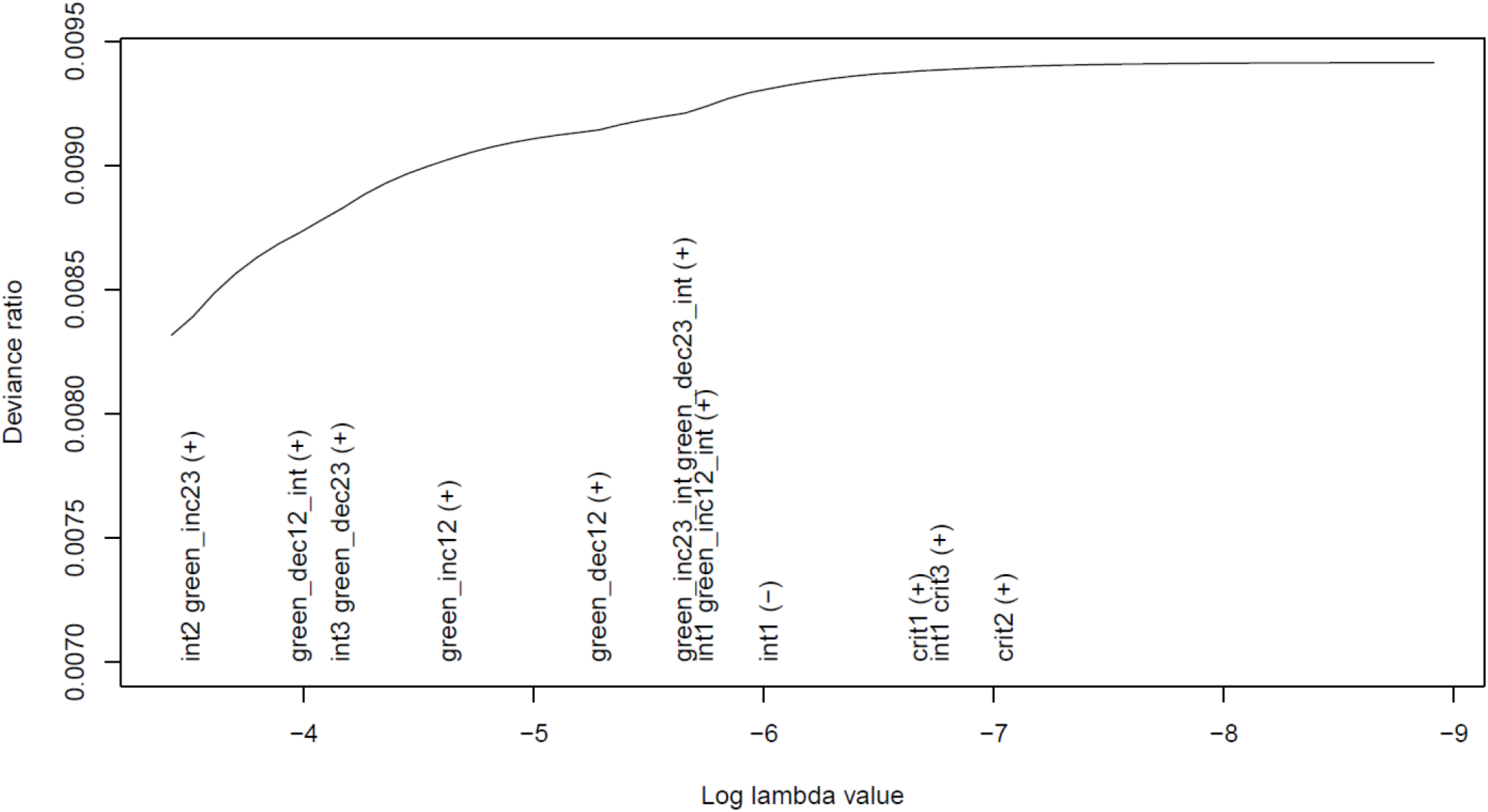
Plot of the lasso model with binary access to green space >5,000m within 300m of home in pregnancy (time 1), at age 4 (time 2) and at age 7 (time 3) as the exposures, BMI at age 7 as the continuous outcome, and highest parental education as the SEP-interaction term (*n* = 6,013). As the variables are added to/removed from the model they appear on the *x*-axis (with “(+)” indicating addition, and “(-)” indicating removal). The covariates/confounders constrained to be included in all models by default do not appear in this plot (SEP, maternal ethnicity, child sex and child age at outcome measurement). There appears to be little association between any of the green space hypotheses and the outcome, with model fit (deviance ratio) barely increasing as variables enter the model. Due to collinearity with the critical period variables, the ‘accumulation’ hypothesis (and hence also its interaction with SEP) was dropped from this model. See figure 1 for a detailed explanation of how to interpret this plot, and table 2 for information on what each of the encoded variables mean.

Similar results were found for the other three outcomes (table S7). For the binary outcome overweight/obese, all SLMCA methods indicated no association between any of the encoded variables and the outcome (figure S5). SBP was a more complex case, as visual inspection indicated at best a marginal increase in model fit as more encoded variables were added (figure S6), the relaxed BIC and the 1 SE cross-validated lassos suggested no association between any of the encoded variables and the outcome, while the relaxed AIC and minimum MSE cross-validated lassos both selected more complex models with 5 and 11 encoded variables, respectively (table S7). Results of the best-fitting relaxed AIC lasso are discussed in detail in table S8, finding that a decrease in access to green space between pregnancy and age 4, and its interaction with SEP, was associated with SBP, with children from lower SEP backgrounds who had a reduction in green space having lower SBP than everyone else. Results for DBP were similar, with some inconsistency again found between the different methods. Visual inspection hinted at a weak association between a decrease in access to green space between pregnancy and age 4 and DBP (figure S7), relaxed BIC and the 1 SE cross-validated lassos indicated no association between any of the encoded variables and the outcome, while the relaxed AIC and minimum MSE cross-validated lassos both selected a decrease in green space between pregnancy and age 4 in their respective best-fitting models (with this decrease in green space associated with lower DBP; table S7).

## Discussion

We have demonstrated how to incorporate multiple exposures and their interaction within a SLCMA, and illustrated three methods of interpreting these results (visual inspection, relaxed lasso via information criteria, and cross-validation). In our simulation study, we observed substantial variation between these different methods in their ability to select the true interaction term. Despite greater ability to detect the interaction term, the relaxed AIC and minimum MSE cross-validation suffered from overfitting and produced numerous false positives, even when the true interaction was null. In contrast, the relaxed BIC and 1 SE cross-validation methods generated few false positives, but frequently did not detect the true interaction term, even when the interaction effect was large (this was especially true for 1 SE cross-validation). The choice of approach may therefore depend on the researcher’s assessment of the risks of under- vs over-fitting and the specific research question; exploratory studies may prefer false positives over false negatives (i.e., over-fitting; and hence use either the relaxed AIC or minimum MSE cross-validated methods), while confirmatory studies may prefer to be more stringent and have greater evidence for an association (i.e., under-fitting; and hence use either relaxed BIC or 1 SE cross-validated methods). Despite these inconsistencies, if different methods give similar results this bolsters confidence in our conclusions. If these methods provide divergent results, then it may not be possible to provide a definitive answer and this ambiguity should be discussed. Nonetheless, as all these methods use the same lasso procedure, the order in which variables are added to the models will be equivalent, so it should be possible to identify the most likely hypotheses which have the strongest association with the outcome (if any).

We also observed considerable variation in the performance of these models to detect the true interaction term given different parameter combinations. Larger sample sizes, centering exposures, and using continuous outcomes may improve the power to detect a true interaction. Models were also more likely to detect an interaction term if collinearity between exposures is lower, and if the interaction effect is larger. These findings may help inform future studies using this methodology, although some of these factors are beyond the researchers’ ability to control.

In our applied example, we found no consistent association between access to green space and child BMI, obesity, SBP or DBP, let alone specific life course trajectories or interactions with SEP. Although some associations between a reduction in access to green space between pregnancy and age 4 and lower blood pressure were reported (tables S5 and S6), these effects were inconsistent, had small effect sizes, and would appear biological implausible (e.g., a reduction in access to green space being associated with lower blood pressure); it is possible that these were a result largely of random noise, especially given how the methods which selected these more complex models – relaxed AIC and minimum MSE cross-validation – are more prone to over-fitting. To the extent that these findings are causal, this suggests that, in this and potentially similar populations at least, access to green space may not improve child BMI or blood pressure.

The applied aspect of this study has several limitations. First, there was lack of variation in exposures over time, which might be expected given that exposures were all based on the child’s home address at three close ages (pregnancy, then when aged 4 and 7 years). Nonetheless, if any association between green space and BMI or blood pressure was present, it should have been visible, even if there was not enough power to detect specific life course trajectories or interactions with SEP. While high collinearity between exposures lowers power to detect a true effect, here we have shown that these methods can be applied even in this challenging scenario; in situations where there is more variation in exposures over time these methods should have even greater ability to uncover the correct life course trajectory. Second, the results from our illustrative example could be biased by measurement error and selection bias and may be explained by residual confounding (Hernán & Robins 2020); such biases need to be considered in any SLCMA studies (Smith *et al*. 2016).

Throughout this paper we have focused on using a standard lasso for SLMCA, rather than the more commonly-used LARS (Smith *et al*. 2015, 2016, 2022). While both approaches are similar, being based on the lasso, the standard lasso approach may have some benefits, especially when considering interactions, including: i) being easier to implement when constraining some covariates to be in the model by default; ii) permitting binary (and other non-continuous) outcomes; and iii) greater flexibility when reporting the results of the best-fitting model (e.g., being able to include main effects of interactions in the reported model, even if not selected in the best-fitting model). Regardless of the SLCMA approach used – lasso or LARS – it is important to check parameter estimates from the final via selective inference to ensure that all encoded variables are associated with the outcome, given the possibility of variables with little-to-no association with the outcome remaining in the final model. For instance, the best-fitting relaxed lasso model via AIC from the applied ALSPAC example with systolic blood pressure as the outcome selected five encoded variables (table S7), yet in the reported model only two were associated with the outcome (table S8).

To improve population health, we need to understand life course trajectories of diseases, and structured life course models are a useful tool for investigating this. We have demonstrated how these structured life course methods can be extended to include interactions between multiple exposures, which should permit a more detailed exploration of how exposures over the life course impact subsequent health. We hope that this paper provides useful guidance for researchers using these methods.

## Data Availability

ALSPAC data access is through a system of managed open access. Information about access to ALSPAC data is given on the ALSPAC website (http://www.bristol.ac.uk/alspac/researchers/access/). The datasets presented in this article are linked to ALSPAC project number B3930, please quote this project number during your application. Simulation and analysis code is openly-available here: https://github.com/djsmith-90/LifeCycle_GreenSpace_CardioOutcomes.

## Ethical Approval

Ethical approval for the study was obtained from the ALSPAC Ethics and Law Committee and the Local Research Ethics Committees. Informed consent for the use of data collected via questionnaires and clinics was obtained from participants following the recommendations of the ALSPAC Ethics and Law Committee at the time.

## Author Contributions

DM-S drafted the initial manuscript and conducted analyses. TD and AE curated and prepared the LifeCycle data for analysis. DM-S, ADACS, DAL and KT contributed to the design of the study. All authors critically reviewed and revised the manuscript.

## Acknowledgements

We are extremely grateful to all the families who took part in this study, the midwives for their help in recruiting them, and the whole ALSPAC team, which includes interviewers, computer and laboratory technicians, clerical workers, research scientists, volunteers, managers, receptionists and nurses. This work was carried out using the computational facilities of the Advanced Computing Research Centre, University of Bristol - http://www.bristol.ac.uk/acrc/.

