## Supplementary Information for "Incorporating interactions into structured life course modelling approaches: A simulation study and applied example of the role of access to green space and socioeconomic position on cardiometabolic health"

*Section S1: Example simulated dataset details*

This section provides details for the data generated in the example simulated dataset as detailed in the 'Worked example' section of the main paper to introduce these structured life course methods when including interaction terms. Equations for the data-generating mechanism are as follows:

$$SEP \sim Bernoulli(0.5)$$

$$green1|SEP \sim Bernoulli(invlogit[\log(0.6) + \log(3) * SEP])$$

$$green2|SEP, green1 \sim Bernoulli(invlogit[\log(0.3) + \log(3) * SEP \ + \log(5) * green1])$$

$$green3|SEP, green2 \sim Bernoulli(invlogit[\log(0.3) + \log(3) * SEP \ + \log(5) * green2])$$

$$BMI|SEP, green1 \sim N(25 + \ -4 * SEP + \ -2 * green3 + 2 * SEP * green3, 3)$$

For the binary 'overweight' outcome, we simply recoded the BMI variable into overweight or not, based on a BMI > 25.

We highlight again that this simulated data is purely to illustrate the logic and application of these structured life-course methods, and should not be taken as a reflection of the real-world patterns or effect sizes of these variables.

*Section S2: Details of data for formal simulation study*

This section provides details for the data generated in formal simulation datasets used in the second section of the paper to explore how well these structured life course methods when including interaction terms perform, given varying parameters, in detecting the true interaction term. The causal relations between the exposures are the same as described in the 'Worked Example' section of the main text. Simulating the outcome BMI depended on the life course hypothesis being assessed, i.e., an interaction between SEP and either critical period at time 3 (as in the worked example), accumulation or change from time 2 to 3. The factors we varied were:

- Sample sizes: comparing *n* = 1,000 vs *n* = 10,000
- Exposure variables: binary vs continuous
- Centering exposure variables: no vs yes
- Collinearity between exposure variables: low vs high
- Outcome variable: binary vs continuous
- Size of interaction term: none vs very small vs small vs moderate vs large vs very large (ranked on a relative scale)
- Life course hypothesis interaction: critical period at time 3 vs accumulation vs change from time 2 to time 3

For all simulations, we used a binary socioeconomic position (SEP) covariate, as described in section S1 of this supplementary material:

$$SEP \sim Bernoulli(0.5)$$

*Exposure variables (access to green space)*

Binary exposure variables with low collinearity (defined as an unadjusted correlation of approximately 0.3 between adjacent exposures) were generated as described in section S1 of this supplementary material:

$$green1|SEP \sim Bernoulli(invlogit[\log(0.6) + \log(3) * SEP])$$

$$green2|SEP, green1 \sim Bernoulli(invlogit[\log(0.3) + \log(3) * SEP \ + \log(5) * green1])$$

$$green3|SEP, green2 \sim Bernoulli(invlogit[\log(0.3) + \log(3) * SEP \ + \log(5) * green2])$$

Binary exposure variables with high collinearity (defined as an unadjusted correlation of approximately 0.9 between adjacent exposures) were generated as follows:

$$green1|SEP \sim Bernoulli(invlogit[\log(0.6) + \log(3) * SEP])$$

$$green2|SEP, green1 \sim Bernoulli(invlogit[\log(0.01) + \log(3) * SEP \ + \log(500) * green1])$$

$$green3|SEP, green2 \sim Bernoulli(invlogit[\log(0.01) + \log(3) * SEP \ + \log(500) * green2])$$

Continuous exposure variables with low collinearity (defined as an unadjusted correlation of approximately 0.5 between adjacent exposures) were generated as follows:

$$green1|SEP \sim N(325 + -50 * SEP, 40])$$

$$green2|SEP, green1 \sim N(240 + -50 * SEP + 0.3 * green1, 40)$$

$$green3|SEP, green2 \sim N(240 + -50 * SEP + 0.3 * green2, 40)$$

Continuous exposure variables with high collinearity (defined as an unadjusted correlation of approximately 0.9 between adjacent exposures) were generated as follows:

$$green1|SEP \sim N(325 + -50 * SEP, 40])$$

$$green2|SEP, green1 \sim N(50 + -50 * SEP + 0.9 * green1, 20)$$

$$green3|SEP, green2 \sim N(240 + -50 * SEP + 0.9 * green2, 20)$$

*Outcome variables (BMI/overweight)*

Continuous outcome variables with binary exposures, where critical period at time 3 and its interaction with SEP caused BMI, were generated as described in section S1 of this supplementary material:

$$BMI|SEP, green1 \sim N(25 + -4 * SEP + -2 * green3 + \gamma * SEP * green3, 3)$$

To assess different strengths of the interaction, we varied the gamma parameter ($\gamma$), at values of 0 (no interaction), 0.5 (very small interaction), 1 (small interaction), 2 (moderate interaction), 3 (large interaction) and 4 (very large interaction).

Continuous outcome variables with continuous exposures, again where critical period at time 3 and its interaction with SEP caused BMI, were generated as follows (the 'green3' parameter was chosen to be 0.02 BMI per unit increase in green space, as the continuous ‘green3’ standard deviation is ~50, and two times this should cover most of the variation in green space distance [0.02 * 50 * 2 = 2], making results broadly comparable to the binary green space effect of 2 BMI units (Gelman 2008)):

$$BMI|SEP, green1 \sim N(28 + -4 * SEP + -0.02 * green3 + \gamma * SEP * green3, 3)$$

To assess different strengths of the interaction, we varied the gamma parameter ($\gamma$), at values of 0 (no interaction), 0.005 (very small interaction), 0.01 (small interaction), 0.02 (moderate interaction), 0.03 (large interaction) and 0.04 (very large interaction).

Continuous outcome variables with binary exposures, where accumulation and its interaction with SEP caused BMI, were generated as follows (with accumulation encoded as in table 2):

$$BMI|SEP, accum \sim N(25 + -4 * SEP + -0.67 * accum + \gamma * SEP * accum, 3)$$

To assess different strengths of the interaction, we varied the gamma parameter ($\gamma$), at values of 0 (no interaction), 0.167 (very small interaction), 0.33 (small interaction), 0.67 (moderate interaction),

1 (large interaction) and 1.33 (very large interaction). These values were chosen to be broadly comparable to the effect sizes in the 'critical period at time 3' scenario.

Continuous outcome variables with continuous exposures, again where accumulation and its interaction with SEP caused BMI, were generated as follows (with accumulation encoded as in table S1):

$$BMI|SEP, accum \sim N(28 + -4 * SEP + -0.02 * accum + \gamma * SEP * accum, 3)$$

To assess different strengths of the interaction, we varied the gamma parameter ($\gamma$), at values of 0 (no interaction), 0.005 (very small interaction), 0.01 (small interaction), 0.02 (moderate interaction), 0.03 (large interaction) and 0.04 (very large interaction).

Continuous outcome variables with binary exposures, where change from time 2 to 3 (using an increase in access to green space between these time points) and its interaction with SEP caused BMI, were generated as follows (with an increase in green space between times 2 and 3 encoded as in table 2):

$$BMI|SEP, change_{inc23} \sim N(25 + -4 * SEP + -2 * change_{inc23} + \gamma * SEP * change_{inc23}, 3)$$

To assess different strengths of the interaction, we varied the gamma parameter ($\gamma$), at values of 0 (no interaction), 0.5 (very small interaction), 1 (small interaction), 2 (moderate interaction), 3 (large interaction) and 4 (very large interaction).

Continuous outcome variables with continuous exposures, again where change from time 2 to 3 and its interaction with SEP caused BMI, were generated as follows (with change in green space between times 2 and 3 encoded as in table S1):

$$BMI|SEP, change_{23} \sim N(25 + -4 * SEP + -0.02 * change_{23} + \gamma * SEP * change_{23}, 3)$$

To assess different strengths of the interaction, we varied the gamma parameter ($\gamma$), at values of 0 (no interaction), 0.005 (very small interaction), 0.01 (small interaction), 0.02 (moderate interaction), 0.03 (large interaction) and 0.04 (very large interaction).

For binary 'overweight' outcome variables, we again simply recoded the BMI variable into overweight or not, based on a BMI > 25.

*Section S3: Additional details of the study population, variable selection and analysis methods for the illustrative ALSPAC example*

### Study population

The Avon Longitudinal Study of Parents and Children (ALSPAC) is a prospective birth cohort study in which pregnant women resident in Avon, UK with expected dates of delivery between 1st April 1991 and 31st December 1992 were invited to take part. The initial number of pregnancies enrolled is 14,541; of these initial pregnancies there were a total of 14,676 fetuses, resulting in 14,062 live births and 13,988 children who were alive at 1 year of age (Boyd *et al.* 2013; Fraser *et al.* 2013). These mothers, their partners, and their index children have been followed with regular questionnaire and clinic assessments since this time. When the oldest children were approximately seven years of age, an attempt was made to bolster the initial sample with eligible cases who had failed to join the study originally. The total sample size for analyses using any data collected after the age of seven is therefore 15,447 pregnancies, resulting in 14,901 children alive at 1 year of age. After removing children or their mothers who had withdrawn consent for their data to be used, the final sample in this analysis consists of 14,845 children.

Please note that the study website contains details of all the data that is available through a fully searchable data dictionary and variable search tool (http://www.bristol.ac.uk/alspac/researchers/our-data/).

### Variable Selection

*Outcome variables*: Our outcome variables were BMI, obesity (BMI ≥ 85th centile [z-score of 1.036] from the age and sex standardised 1990 UK growth reference charts (Cole *et al.* 1995)) and systolic and diastolic blood pressure assessed at a follow-up research clinic when the study children were approximately 7 years of age.

*Exposure variables:* ALSPAC is part of the LifeCycle project EU Child Cohort Network, an initiative to harmonise data across a network of European birth/childhood cohort studies to investigate how early-life stressors impact health across the lifespan (Jaddoe *et al.* 2020). Part of this harmonisation process includes repeated measures of the built environments, such as access to green space, which will be used as our exposures for the present study. We used the LifeCycle variable 'whether there is a green space >5,000 $m^2$ within 300 meters of the geocoded family home address' as our binary access to green space exposure. This definition is frequently used by policy makers to define 'residential proximity to a major green space', and is approximately a 5 minute walk (https://www.euro.who.int/__data/assets/pdf_file/0010/342289/Urban-Green-Spaces_EN_WHO_web3.pdf). The European 'Urban Atlas' database, produced by the European Environmental Protection Agency (2010), based on green space data from 2006 was used to determine the location of green spaces, its size, and its distance from the family home (https://www.eea.europa.eu/data-and-maps/data/urban-atlas). The Barcelona Institute for Global Health (ISGlobal) linked the Urban Atlas green space layers with family geocoded addresses in a centralised Geographic Information System (GIS) environment to generate the green space exposure variable and harmonise it across cohort studies (de Castro Pascual *et al.* 2021). The ALSPAC Data team then received and prepared the data for release. In this study, we used three measures of this binary ‘access to green space’ exposure; in pregnancy (time 1), when the child was aged 4 years of age (time 2), and when the child was aged 7 years of age (time 3).

*SEP Confounder/Interaction*: We used highest parental education as our SEP exposure, with A-levels (optional qualifications taken at approximately age 18 year) or higher coded as ‘high’ and O-level (qualifications taken at approximately age 16 years) or lower coded as ‘low’. This variable was measured in pregnancy.

*Other confounders/covariates*: Based on our hypothesised DAG (figure S2), the only factor that may confound these associations is ethnicity, so this variable was included in all analyses (coded as White vs other than White). Other covariates which are not necessarily confounders, but may help predict the outcomes and remove residual variation, which were included in analyses were the sex of the child and age of the child at the time of outcome measurement.

**Analysis**

We followed the same SLCMA detailed above for the simulated data, analysing the data using the same three methods: visual inspection, a ‘relaxed lasso’ approach (using both AIC and BIC to identify the best-fitting models), and cross-validated lasso (using both the minimum prediction error and within 1 standard error of this value to select the best-fitting models). Each set of SLCMA models was repeated for each of the four outcomes, using a linear model for continuous outcomes (BMI, SBP and DBP) and a logistic model for binary outcomes (overweight/obesity). As all exposures were binary, hypotheses were encoded as described in table 2. All exposures were mean-centered in an attempt to reduce the correlation between the exposure and the interaction term and increase power. To avoid collinearity between encoded variables, variables with a correlation coefficient greater than $r = 0.9$ with other variables were removed from the set of hypotheses to be tested. In all models this resulted in the ‘accumulation’ hypothesis (and hence also the ‘accumulation interaction’ term) being omitted due to collinearity with the critical period variables.

*Table S1:* Hypotheses encoded in our simulation study where green space exposures were continuous variables. The variables are: green1 (distance to green space at first time point), green2 (distance to green space at second time point), green3 (distance to green space at third time point) and high_sep (binary marker for high socioeconomic position). The confounder 'high_sep' is included in all models. Simple models can be encoded in just one variable, while compound hypotheses combine two or more simple hypotheses. SEP = socioeconomic position.

| **Hypothesis** | **Encoding** | **Type** |
|---|---|---|
| Critical period (1) | crit1 = green1 | Simple |
| Critical period (1) only if high SEP | int1 = high_sep * green1 | Simple |
| Critical period (1) with SEP interaction | crit1 + int1 | Compound |
| Critical period (2) | crit2 = green2 | Simple |
| Critical period (2) only if high SEP | int2 = high_sep * green2 | Simple |
| Critical period (2) with SEP interaction | crit2 + int2 | Compound |
| Critical period (3) | crit3 = green3 | Simple |
| Critical period (3) only if high SEP | int3 = high_sep * green3 | Simple |
| Critical period (3) with SEP interaction | crit3 + int3 | Compound |
| Accumulation | accumulation = (green1 + green2 + green3) / 3 | Simple |
| Accumulation only if high SEP | int_accum = high_sep * accumulation | Simple |
| Accumulation with SEP interaction | accumulation + int_accum | Compound |
| Sensitive period (1) | crit1 + accumulation | Compound |
| Sensitive period (2) | crit2 + accumulation | Compound |
| Sensitive period (3) | crit3 + accumulation | Compound |
| Change in green space from time 1 to 2 | green_ch12 = green2 – green1 | Simple |
| Change in green space from time 1 to 2 only if high SEP | green_ch12_int = high_sep * green_ch12 | Simple |
| Change in green space from time 1 to 2 with SEP interaction | green_ch12 + green_ch12_int | Compound |
| Change in green space from time 2 to 3 | green_ch23 = green3 – green2 | Simple |
| Change in green space from time 2 to 3 only if high SEP | green_ch23_int = high_sep * green_ch23 | Simple |
| Change in green space from 2 to 3 with SEP interaction | green_ch23 + green_ch23_int | Compound |

*Table S2:* Summary of overall simulation results for each SLCMA method, each interaction strength, and each life course interaction model, split by continuous and binary outcomes (*n* simulations = 16; *n* iterations per simulation = 1,000). Results show the percent of simulations in which the correct interaction term was selected. Note that in the rows for interaction strength of 'None', technically this is not the 'correct' interaction as no interaction was present (i.e., 8.4% of simulations using the AIC relaxed lasso method with a continuous outcome included the interaction term for the critical period at time 3 model, even though no interaction was simulated). For full details on the simulation parameters, see section S2 of the supplementary information. Note that models with binary outcomes and an interaction strength of 'none' were more likely to include the interaction term compared to an interaction strength of 'very small'; this is because the dichotomisation of the continuous BMI outcome made the interaction effect slightly negative, compared to the original interaction term with the continuous outcome (e.g., where the true interaction is null for the continuous outcome it is slightly negative for the binary outcome, while where the true interaction is very small for the continuous outcome, it is practically null for the binary outcome). SE = Standard error; MSE = Mean-squared error.

| **Interaction Strength** | **Life course interaction model** | **Continuous outcome** | | | | **Binary outcome** | | | |
|---|---|---|---|---|---|---|---|---|---|
| | | **Relaxed lasso – AIC** | **Relaxed lasso – BIC** | **Cross-validated lasso – 1 SE of min. MSE** | **Cross-validated lasso – min. MSE** | **Relaxed lasso – AIC** | **Relaxed lasso – BIC** | **Cross-validated lasso – 1 SE of min. MSE** | **Cross-validated lasso – min. MSE** |
| None | Critical period at time 3 | 8.4% | 1.1% | 1.5% | 19.5% | 23.1% | 3.5% | 0.0% | 29.1% |
| | Accumulation | 3.7% | 1.1% | 0.6% | 9.2% | 6.6% | 0.6% | 0.0% | 10.6% |
| | Change from time 2 to 3 | 17.6% | 3.0% | 0.3% | 34.5% | 24.7% | 2.3% | 0.0% | 31.8% |
| Very small | Critical period at time 3 | 42.9% | 9.7% | 0.1% | 52.2% | 7.4% | 0.3% | 0.0% | 11.9% |
| | Accumulation | 10.3% | 0.9% | 0.0% | 26.6% | 1.5% | 0.1% | 0.0% | 3.2% |
| | Change from time 2 to 3 | 41.3% | 13.2% | 0.0% | 44.8% | 11.3% | 0.4% | 0.0% | 13.9% |
| Small | Critical period at time 3 | 67.4% | 45.2% | 0.6% | 73.6% | 41.5% | 10.0% | 0.0% | 47.3% |
| | Accumulation | 30.2% | 13.5% | 0.1% | 52.6% | 8.2% | 0.7% | 0.0% | 16.7% |
| | Change from time 2 to 3 | 61.4% | 36.2% | 0.7% | 60.8% | 32.3% | 9.1% | 0.0% | 31.3% |
| Moderate | Critical period at time 3 | 92.6% | 77.1% | 42.7% | 97.5% | 78.0% | 52.9% | 14.4% | 81.2% |
| | Accumulation | 56.2% | 38.6% | 17.1% | 73.5% | 38.9% | 23.2% | 2.0% | 51.8% |
| | Change from time 2 to 3 | 82.4% | 59.9% | 24.7% | 79.8% | 64.2% | 41.0% | 6.4% | 60.9% |
| Large | Critical period at time 3 | 98.4% | 94.5% | 71.6% | 99.9% | 93.8% | 75.1% | 51.1% | 96.5% |
| | Accumulation | 72.8% | 58.2% | 44.0% | 83.6% | 57.6% | 40.5% | 27.2% | 66.3% |
| | Change from time 2 to 3 | 90.5% | 73.1% | 40.9% | 88.0% | 81.7% | 59.1% | 28.3% | 76.7% |
| Very large | Critical period at time 3 | 99.8% | 99.0% | 92.7% | 100.0% | 97.8% | 88.0% | 67.9% | 98.9% |
| | Accumulation | 80.4% | 73.3% | 59.2% | 84.2% | 67.7% | 53.4% | 36.5% | 75.5% |
| | Change from time 2 to 3 | 94.9% | 86.3% | 56.8% | 92.4% | 89.4% | 74.6% | 41.0% | 85.5% |

*Table S3:* Summary of simulation results for each SLMCA method and each life course interaction models, under the 'moderate interaction' scenario, with results separated by each of the simulation parameters varied and by continuous vs binary outcomes (*n* simulations = 16; *n* iterations per simulation = 1,000). Results show the percent of simulations in which the correct interaction term was selected (as there were 16 simulations for each type of outcome variable, each row here contains the average over 8 simulations). For full details on the simulation parameters, see section S2 of the supplementary information. SE = Standard error; MSE = Mean-squared error.

| **Simulation parameter** | **Life course interaction model** | **Relaxed lasso – AIC** | **Relaxed lasso – BIC** | **Cross-validated lasso – 1 SE of min. MSE** | **Cross-validated lasso – min. MSE** |
|---|---|---|---|---|---|
| *Sample size – Continuous outcome* | | | | | |
| 1,000 | Critical period at time 3 | 86.0% | 55.1% | 4.4% | 95.0% |
| | Accumulation | 29.3% | 8.1% | 0.4% | 52.7% |
| | Change from time 2 to 3 | 69.4% | 41.5% | 4.7% | 66.8% |
| 10,000 | Critical period at time 3 | 99.2% | 99.0% | 81.0% | 100.0% |
| | Accumulation | 83.2% | 69.1% | 33.7% | 94.4% |
| | Change from time 2 to 3 | 95.3% | 78.2% | 44.7% | 92.8% |
| *Sample size – Binary outcome* | | | | | |
| 1,000 | Critical period at time 3 | 58.0% | 12.6% | 0.5% | 62.5% |
| | Accumulation | 12.4% | 1.2% | 0.1% | 20.6% |
| | Change from time 2 to 3 | 44.1% | 14.5% | 0.4% | 40.5% |
| 10,000 | Critical period at time 3 | 98.1% | 93.3% | 28.4% | 100.0% |
| | Accumulation | 65.4% | 45.1% | 3.9% | 83.1% |
| | Change from time 2 to 3 | 84.2% | 67.4% | 12.3% | 81.2% |
| *Exposure – Continuous outcome* | | | | | |
| Binary | Critical period at time 3 | 93.9% | 82.6% | 52.9% | 97.8% |
| | Accumulation | 68.0% | 49.2% | 30.4% | 78.3% |
| | Change from time 2 to 3 | 73.6% | 42.6% | 19.9% | 69.3% |
| Continuous | Critical period at time 3 | 91.3% | 71.6% | 32.5% | 97.2% |
| | Accumulation | 44.5% | 27.9% | 3.7% | 68.7% |
| | Change from time 2 to 3 | 91.2% | 77.1% | 29.4% | 90.2% |
| *Exposure – Binary outcome* | | | | | |
| Binary | Critical period at time 3 | 78.9% | 56.6% | 16.5% | 80.3% |
| | Accumulation | 45.4% | 29.8% | 3.3% | 54.5% |
| | Change from time 2 to 3 | 49.4% | 27.7% | 0.8% | 43.2% |
| Continuous | Critical period at time 3 | 77.2% | 49.3% | 12.4% | 82.2% |
| | Accumulation | 32.3% | 16.6% | 0.7% | 49.1% |
| | Change from time 2 to 3 | 78.9% | 54.2% | 11.9% | 78.5% |
| *Collinearity of exposures – Continuous outcome* | | | | | |
| Low | Critical period at time 3 | 97.8% | 81.6% | 43.9% | 98.7% |
| | Accumulation | 69.1% | 51.1% | 12.7% | 80.8% |
| | Change from time 2 to 3 | 98.5% | 88.1% | 49.1% | 98.1% |
| High | Critical period at time 3 | 87.4% | 72.5% | 41.5% | 96.4% |
| | Accumulation | 43.4% | 26.1% | 21.4% | 66.3% |

| | | | | | |
|---|---|---|---|---|---|
| | Change from time 2 to 3 | 66.3% | 31.7% | 0.2% | 61.5% |
| *Collinearity of exposures – Binary outcome* | | | | | |
| Low | Critical period at time 3 | 83.0% | 57.3% | 16.1% | 83.1% |
| | Accumulation | 51.8% | 33.7% | 0.5% | 58.1% |
| | Change from time 2 to 3 | 83.4% | 63.3% | 12.7% | 81.5% |
| High | Critical period at time 3 | 73.0% | 48.5% | 12.8% | 79.4% |
| | Accumulation | 26.0% | 12.7% | 3.4% | 45.5% |
| | Change from time 2 to 3 | 44.9% | 18.7% | 0.0% | 40.3% |
| *Encoded variables centered – Continuous outcome* | | | | | |
| No | Critical period at time 3 | 94.9% | 76.8% | 42.6% | 97.7% |
| | Accumulation | 47.8% | 32.4% | 14.2% | 69.6% |
| | Change from time 2 to 3 | 83.0% | 60.3% | 24.5% | 80.1% |
| Yes | Critical period at time 3 | 90.3% | 77.3% | 42.8% | 97.3% |
| | Accumulation | 64.7% | 44.8% | 20.0% | 77.5% |
| | Change from time 2 to 3 | 81.7% | 59.4% | 24.9% | 79.5% |
| *Encoded variables centered – Binary outcome* | | | | | |
| No | Critical period at time 3 | 79.4% | 54.4% | 12.5% | 80.2% |
| | Accumulation | 29.8% | 15.8% | 1.3% | 44.9% |
| | Change from time 2 to 3 | 65.2% | 41.2% | 6.4% | 61.5% |
| Yes | Critical period at time 3 | 76.7% | 51.4% | 16.4% | 82.3% |
| | Accumulation | 48.0% | 30.5% | 2.6% | 58.7% |
| | Change from time 2 to 3 | 63.1% | 40.7% | 6.3% | 60.2% |

*Table S4*: Descriptive statistics of the ALSPAC binary 'access to green space' exposure (*n* = 14,845). Only participants with data at all three time-points have been included; if only one or two time-points had data, they have been coded as missing.

| **Time point** | **Coding** | **N (%)** |
|---|---|---|
| Access to green space >5,000m within 300m of home in pregnancy | Yes | 8,961 (75.9%) |
| | No | 2,838 (25.1%) |
| Access to green space >5,000m within 300m of home at age 4 | Yes | 9,024 (76.5%) |
| | No | 2,775 (23.5%) |
| Access to green space >5,000m within 300m of home at age 7 | Yes | 9,093 (77.1%) |
| | No | 2,706 (22.9%) |
| Missing (%) | | 3,046 (20.5%) |

*Table S5:* Variation in binary access to green space >5,000m within 300m of home over each of the three ALSPAC time-points (pregnancy, age 4 and age 7; *n* = 11,799).

| **Access to green space in pregnancy** | **Access to green space at age 4** | **Access to green space at age 7** | **N (%)** |
|---|---|---|---|
| No | No | No | 2,054 (17.4%) |
| Yes | No | No | 401 (3.4%) |
| No | Yes | No | 22 (0.2%) |
| No | No | Yes | 242 (2.1%) |
| Yes | Yes | No | 229 (1.9%) |
| Yes | No | Yes | 78 (0.7%) |
| No | Yes | Yes | 520 (4.4%) |
| Yes | Yes | Yes | 8,253 (69.9%) |

*Table S6*: Descriptive statistics of ALSPAC outcomes and covariates (*n* = 14,845). N (%) for categorical variables; mean (standard deviation [SD]) for continuous variables.

| **Variable** | | **N (%); Mean (SD)** |
|---|---|---|
| Body mass index (BMI; kg/m$^2$) | | 16.3 (2.1) |
| Missing (%) | | 6,662 (44.9%) |
| Overweight/obese | Yes | 1,476 (18.0%) |
| | No | 6,707 (82.0%) |
| Missing (%) | | 6,662 (44.9%) |
| Systolic blood pressure (mmHg) | | 99.0 (9.2) |
| Missing (%) | | 6,784 (45.7%) |
| Diastolic blood pressure (mmHg) | | 56.5 (6.7) |
| Missing (%) | | 6,786 (45.7%) |
| SEP – Parental education | Higher (A-level or higher) | 6,570 (55.0%) |
| | Lower (O-level or lower) | 5,375 (45.0%) |
| Missing (%) | | 2,900 (19.5%) |
| Sex of child at birth | Female | 7,269 (49.0%) |
| | Male | 7,576 (51.0%) |
| Missing (%) | | 0 (0%) |
| Age of child at outcome measurement (months) | | 89.0 (3.8) |
| Missing (%) | | 6,583 (44.3%) |
| Maternal ethnicity | White | 11,966 (97.4%) |
| | Other than White | 323 (2.6%) |
| Missing (%) | | 2,556 (17.2%) |

*Table S7:* Results of the structured life course models using ALSPAC data. The exposure is a binary 'access to green space' variable measured at three time points (pregnancy, when the study child was age 4 years, and when the study child was aged 7 years), and the interaction term is socioeconomic position (SEP; highest parental education, as measured in pregnancy). BMI = body mass index; MSE = Mean-squared error; SE = Standard error.

| **Outcome** | **Visual inspection** | **Relaxed lasso** | | **Cross-validated lasso** | |
|---|---|---|---|---|---|
| | | *Minimum AIC model* | *Minimum BIC model* | *Model within 1 SE of minimum MSE* | *Minimum MSE model* |
| *BMI* (n = *6,013)* | No obvious associations/ minimal improvement in model fit (figure 4) | No variables beyond baseline confounders/ covariates | No variables beyond baseline confounders/ covariates | No variables beyond baseline confounders/ covariates | No variables beyond baseline confounders/ covariates |
| *Overweight/ obese (*n = *6,013)* | No obvious associations/ minimal improvement in model fit (figure S5) | No variables beyond baseline confounders/ covariates | No variables beyond baseline confounders/ covariates | No variables beyond baseline confounders/ covariates | No variables beyond baseline confounders/ covariates |
| *Systolic blood pressure (*n = *5,918)* | No obvious associations/ minimal improvement in model fit (figure S6) | Five encoded variables included beyond baseline confounders/ covariates (see table S8 for more details) | No variables beyond baseline confounders/ covariates | No variables beyond baseline confounders/ covariates | Eleven encoded variables included beyond baseline confounders/ covariates[a] |
| *Diastolic blood pressure (*n = *5,918)* | Perhaps weak evidence for a decrease in green space between pregnancy and age 4 associated with outcome (figure S7) | One encoded variable included beyond baseline confounders/ covariates (decrease in green space between pregnancy and age 4)[b] | No variables beyond baseline confounders/ covariates | No variables beyond baseline confounders/ covariates | One encoded variable included beyond baseline confounders/ covariates (decrease in green space between pregnancy and age 4)[b] |

[a] Results not displayed here, but they are very similar to the relaxed AIC results for systolic blood pressure described in table S8 (i.e., of the encoded variables, only a decrease in green space between pregnancy and age 4, and its interaction with socioeconomic position, were associated with the outcome).

[b] Post-selection inference suggested that a decrease in green space between pregnancy and age 4 was associated with a reduction in diastolic blood pressure ($b$ = -1.209, SE = 0.443, 95% confidence interval = -2.084 to -0.285, $p$ = 0.006).

*Table S8:* Assessing the ALSPAC result in detail where the relaxed AIC lasso for systolic blood pressure outcome found 5 encoded variables (beyond the baseline confounders/covariates) in the best-fitting model. These encoded variables were: 'int3' (interaction between SEP and critical period at age 7); 'green_dec12' (decrease in green space between pregnancy and age 4); 'green_dec12_int' (interaction between SEP and decrease in green space between pregnancy and age 4); 'green_inc_23' (interaction between SEP and increase in green space between ages 4 and 7) and; 'green_dec23' (decrease in green space between age 4 and 7). To explore these results in more detail, we performed post-selection inference to estimate unbiased confidence intervals and *p*-values for this model (for more details on this method, see: (Tibshirani *et al.* 2016; Smith *et al.* 2022)). As some of these selected encoded variables were interaction terms without a main effect, we compared results both excluding and including these additional main effects. These results suggest that the only robust effect is that of decrease in green space from pregnancy to age 4 and its interaction with SEP. These models indicate that a decrease in access to green space between pregnancy and age 4 is associated with lower systolic blood pressure, but only among those from low SEP backgrounds (with systolic blood pressure being approximately equivalent for those who did not experience a decrease in access to green space between this time, and for those who did experience a decrease but came from high SEP backgrounds). This association is counter-intuitive, given that a decrease in access to green space may be expected to worsen – not improve – blood pressure. Given this, the complexity of the model, the minimal improvement in model fit (figure S6), the fact that other SLCMA models with this outcome did not detect this association (e.g., the relaxed BIC and cross-validated 1SE models), and the lack of similar effects in other literature, this would perhaps suggest that this result may be largely due to random noise in the data rather than a meaningful biological effect. SE = Standard error; CI = Confidence interval.

| **Variable** | **Without additional main effects (i.e., the model selected by the lasso)** | | | **With additional main effects where only interaction terms were selected by the lasso** | | |
|---|---|---|---|---|---|---|
| | *Beta (SE)* | *95% CI* | p-*value* | *Beta (SE)* | *95% CI* | p-*value* |
| Age (months) | 0.286 (-0.053) | 0.181 to 0.390 | <0.001 | 0.285 (-0.053) | 0.181 to 0.390 | <0.001 |
| Sex (1 = male) | -0.340 (0.236) | -0.803 to 0.355 | 0.236 | -0.337 (0.236) | -0.804 to 0.364 | 0.154 |
| Ethnicity (1 = White) | 0.527 (0.868) | -4.878 to 2.151 | 0.868 | 0.520 (0.868) | -4.967 to 2.134 | 0.550 |
| SEP (1 = High) | -1.164 (0.244) | -1.647 to -0.685 | <0.001 | -1.169 (0.244) | -1.652 to -0.690 | <0.001 |
| Critical period at age 7 | - | - | - | -0.336 (0.502) | -1.307 to 2.527 | 0.504 |
| Critical period at age 7 with SEP interaction | 0.867 (0.384) | 0.004 to 1.627 | 0.024 | 1.188 (0.616) | -1.604 to 2.383 | 0.616 |
| Decrease in green space from pregnancy to age 4 | -2.807 (0.900) | -4.583 to -0.983 | 0.002 | -2.844 (0.993) | -5.175 to -0.456 | 0.011 |
| Decrease in green space from pregnancy to age 4 with SEP interaction | 3.708 (1.258) | 1.116 to 6.198 | 0.003 | 3.732 (1.321) | 0.984 to 6.482 | 0.005 |
| Increase in green space from age 4 to 7 | - | - | - | -0.741 (1.173) | -2.955 to 6.306 | 0.528 |
| Increase in green space from age 4 to 7 with SEP interaction | 0.929 (0.852) | -2.200 to 2.583 | 0.852 | 1.671 (1.449) | -5.485 to 4.450 | 0.352 |
| Decrease in green space from age 4 to 7 | 1.113 (0.787) | -1.228 to 2.657 | 0.787 | 0.968 (0.812) | -1.799 to 2.678 | 0.234 |

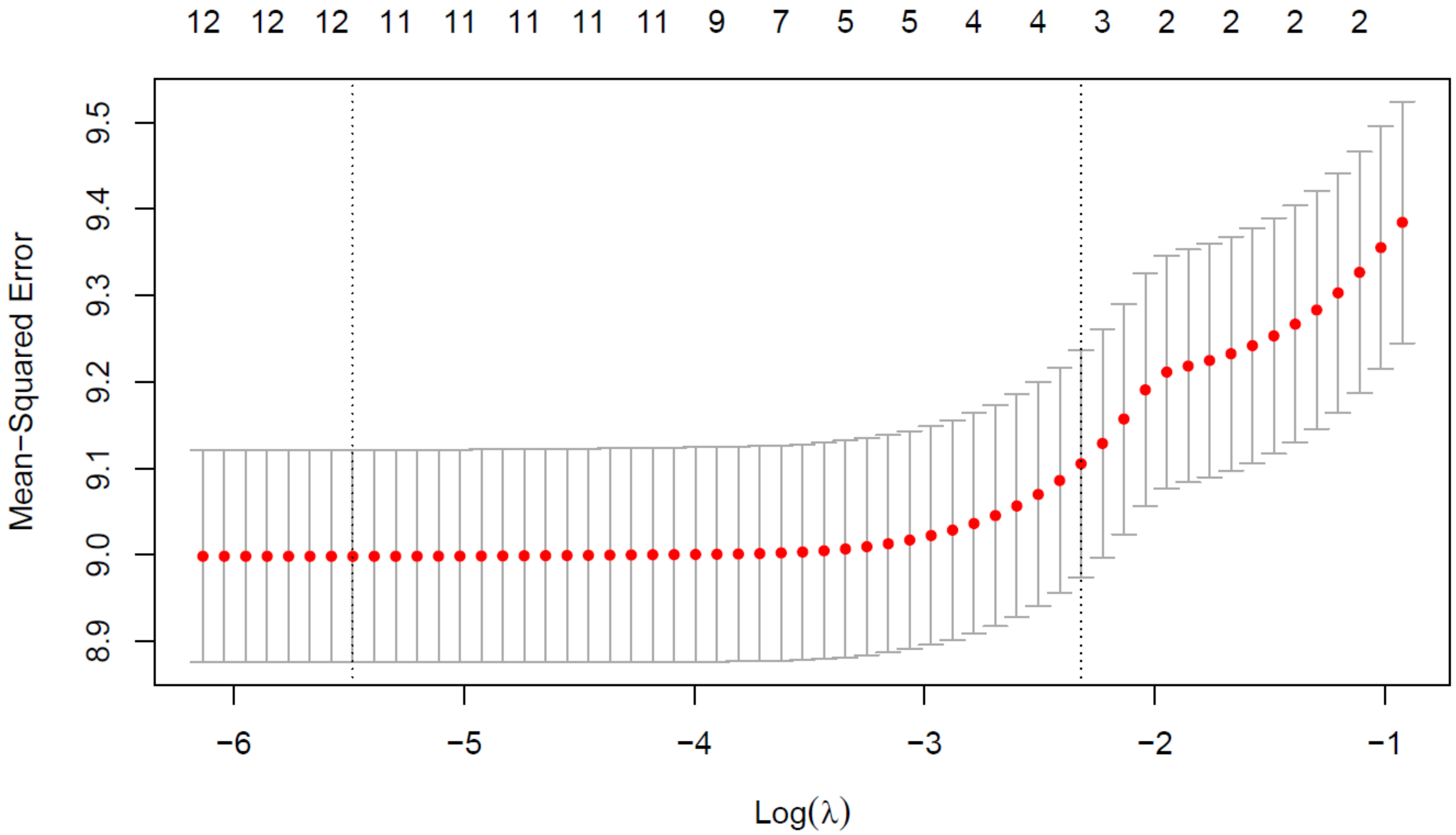

*Figure S1:* Example of the output from the cross-validated lasso using the simulated example dataset. The log-lambda value is displayed on the *x*-axis, with decreasing lambda values going from right to left. The mean-squared error (*y*-axis) is a measure of out-of-sample prediction error, based on splitting the data into *k* equal portions (here, *k* = 10) and calculating the mean prediction error for each lambda value. The left dotted vertical line denotes the lambda value which minimises the prediction error, while the right dotted vertical line is the model where the prediction error is no more than one standard error above the minimum mean-squared error value. The number of parameters included in each model is given above the plot. In the one standard error model, there are three variables included: the SEP variable (‘high_sep’; included by default), the most recent critical period variable (‘crit3’), and the interaction between SEP and the most recent critical period (‘int3’), just as we simulated. The model with the lowest prediction error contains these three parameters, plus nine additional encoded hypotheses.

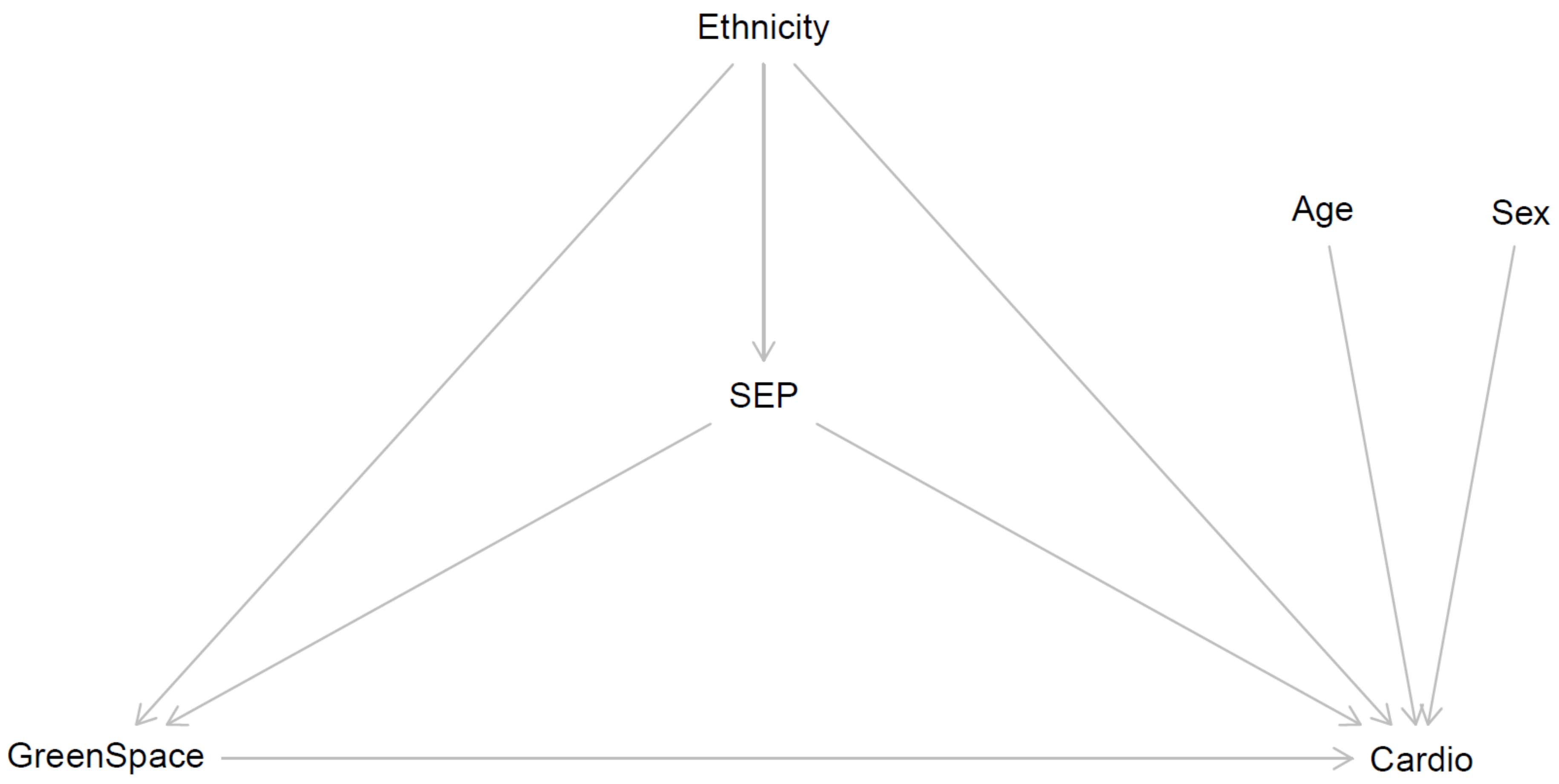

*Figure S2:* Hypothesised Directed Acyclic Graph (DAG) for the ALSPAC data. Note that for simplicity only one 'green space' node, representing all three measured time-points, has been included here. We are also assuming that there may be a potential interaction between SEP and access to green space on cardiometabolic outcomes, which is not explicitly represented in this DAG. SEP = Socioeconomic position.

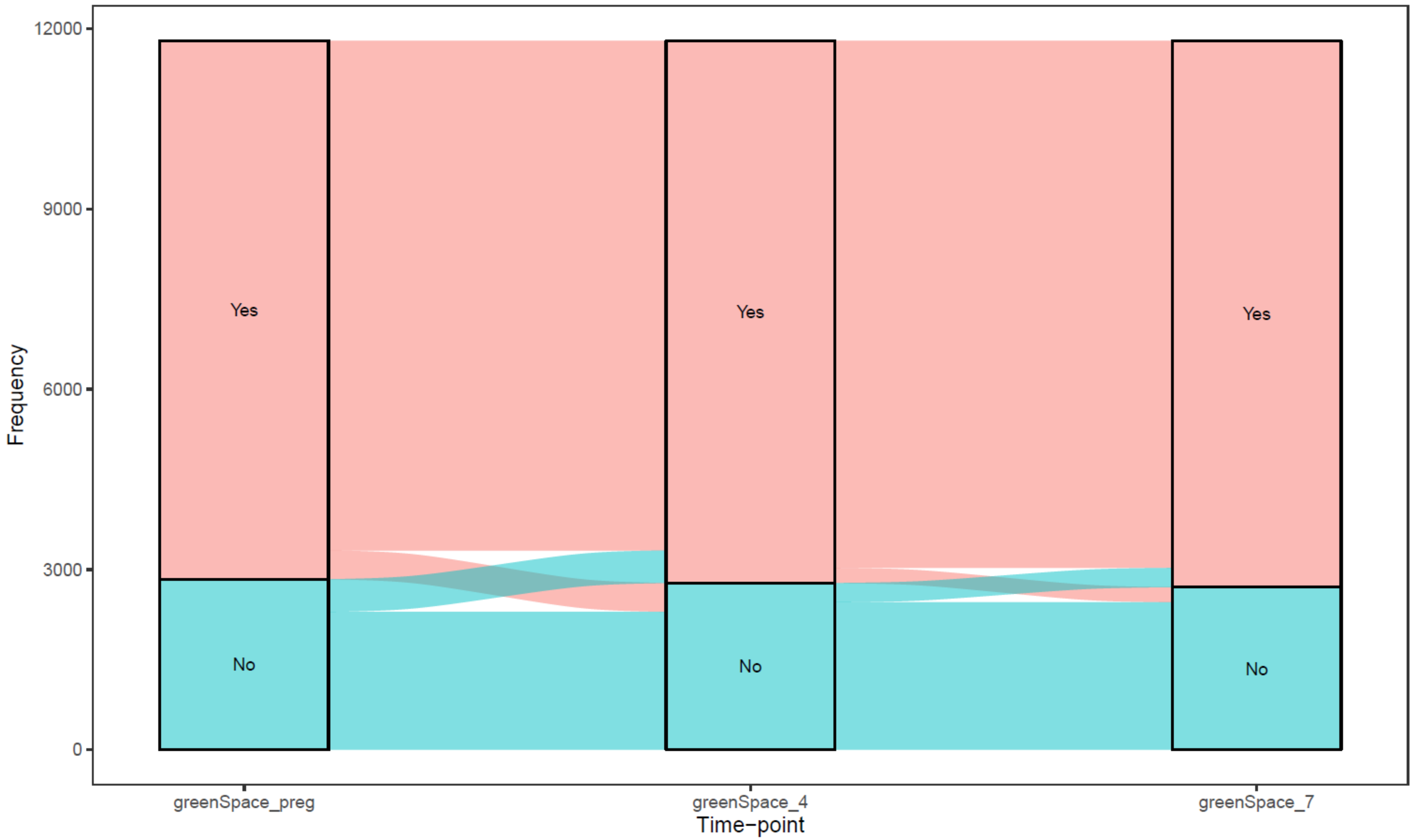

*Figure S3:* Sankey plot displaying the change in binary access to green space (>5,000 m$^2$ within 300 meters of home address) at each time-point ($n$ = 11,799).

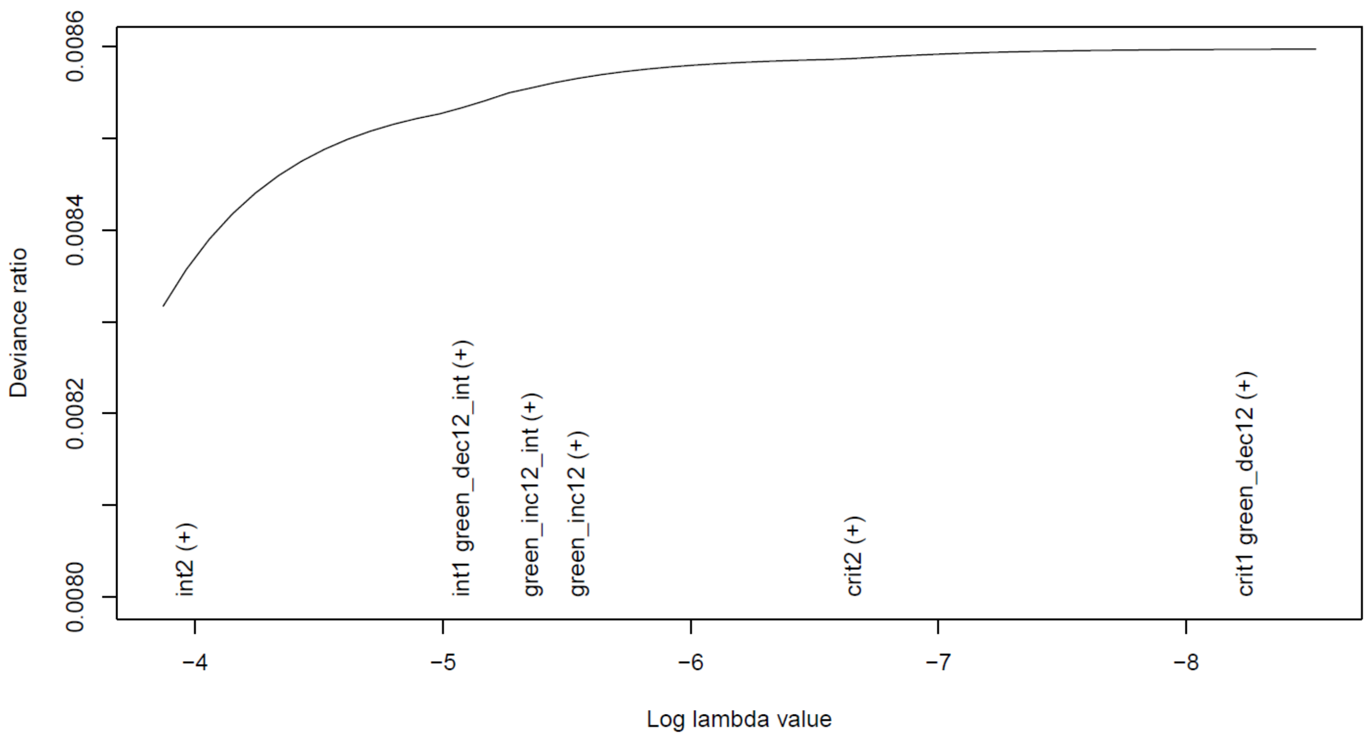

*Figure S4:* Plot of the lasso model with binary access to green space >5,000m within 300m of home in pregnancy (time 1) and at age 7 (time 2) as the exposures, BMI at age 7 as the continuous outcome, and highest parental education as the SEP-interaction term (*n* = 6,013). As the variables are added to/removed from the model they appear on the *x*-axis (with "(+)" indicating addition, and "(-)" indicating removal). The covariates/confounders constrained to be included in all models by default do not appear in this plot (SEP, maternal ethnicity, child sex and child age at outcome measurement). Unlike figure 4 in the main text, this model only includes exposures from pregnancy and age 7 (i.e., excluding age 4); results are qualitatively identical, however (i.e., little/no association between access to green space and child BMI; even though 'int2' [interaction between SEP and critical period at age 7]] was entered before all other variables, the improvement in model fit is minimal, ~0.02 percentage point improvement in the deviance ratio before the next variables were added). This result was confirmed by the relaxed lasso (AIC and BIC) and cross-validated lasso (minimum MSE and 1SE of MSE), which all selected the covariate-only model as the best fit to the data. Due to collinearity with the critical period variables, the 'accumulation' hypothesis (and hence also its interaction with SEP) was dropped from this model. See figure 1 for a detailed explanation of how to interpret this plot, and table 2 for information on what each of the encoded variables mean.

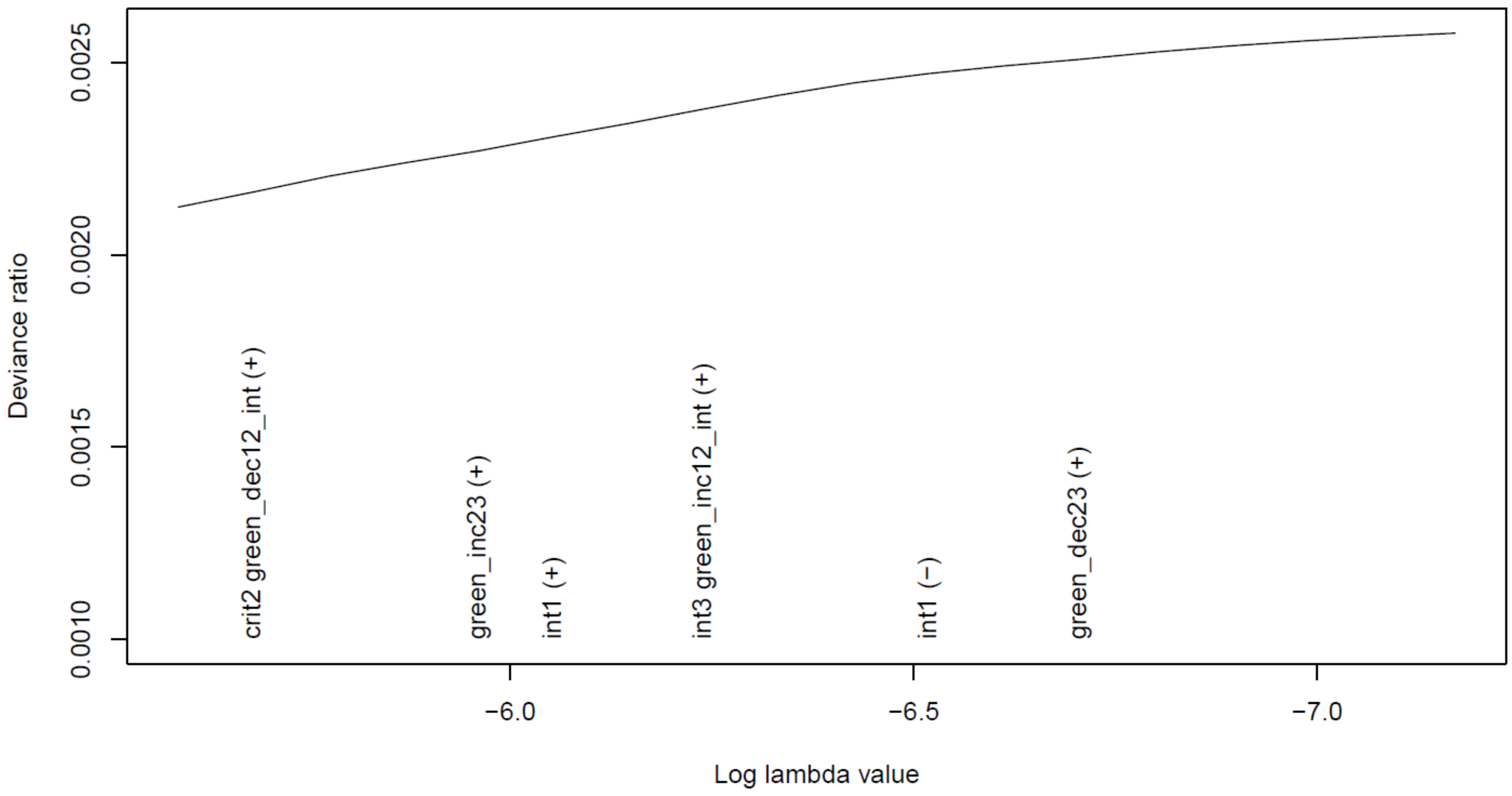

*Figure S5:* Plot of the lasso model with binary access to green space >5,000m within 300m of home in pregnancy (time 1), at age 4 (time 2) and at age 7 (time 3) as the exposures, overweight/obese at age 7 as the binary outcome, and highest parental education as the SEP-interaction term (*n* = 6,013). As the variables are added to/removed from the model they appear on the *x*-axis (with "(+)" indicating addition, and "(-)" indicating removal). The covariates/confounders constrained to be included in all models by default do not appear in this plot (SEP, maternal ethnicity, child sex and child age at outcome measurement). There appears to be little association between any of the green space hypotheses and the outcome, with model fit (deviance ratio) barely increasing as variables enter the model (see table S7 for a more formal test using relaxed and cross-validated lasso approaches). Due to collinearity with the critical period variables, the 'accumulation' hypothesis (and hence also its interaction with SEP) was dropped from this model. See figure 1 for a detailed explanation of how to interpret this plot, and table 2 for information on what each of the encoded variables mean.

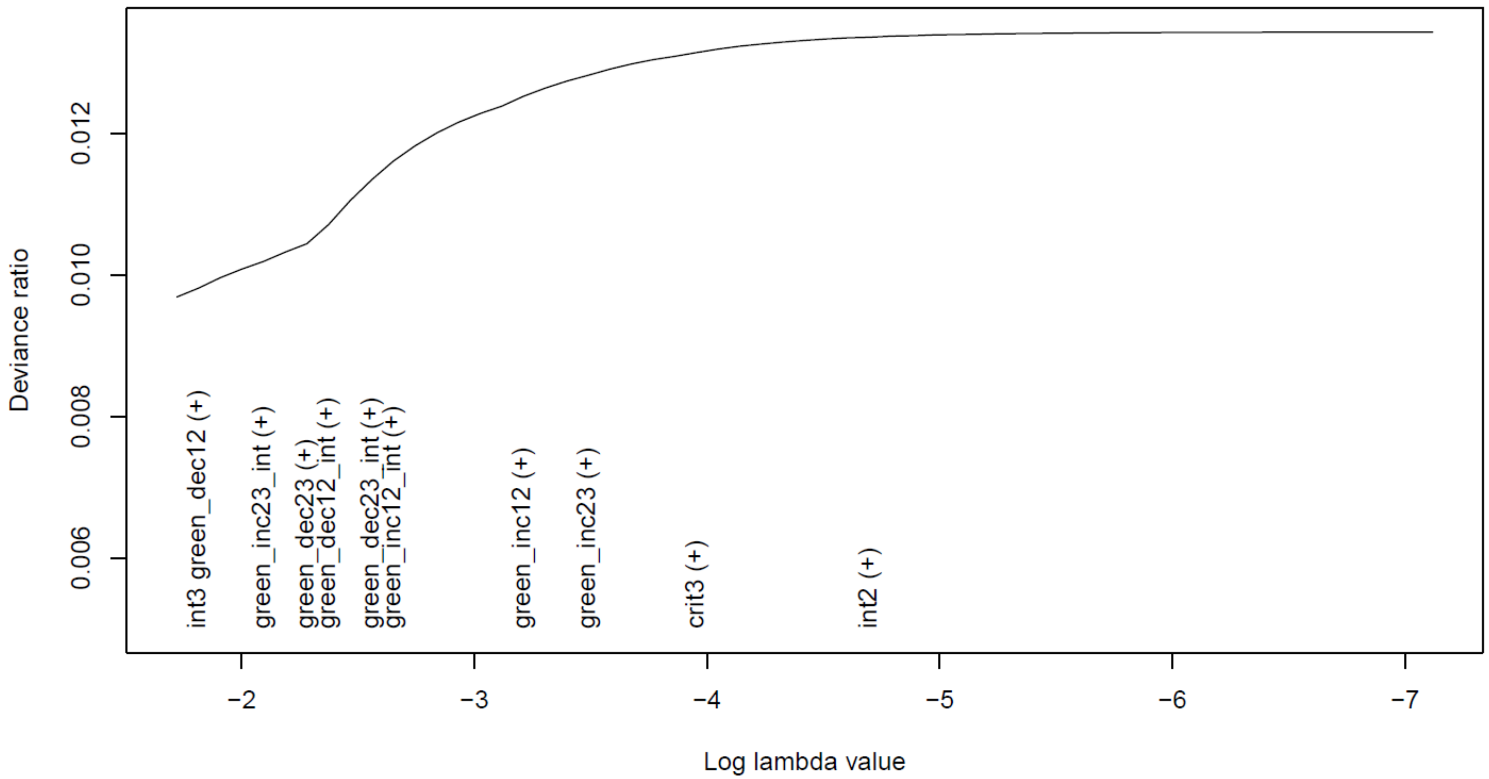

*Figure S6:* Plot of the lasso model with binary access to green space >5,000m within 300m of home in pregnancy (time 1), at age 4 (time 2) and at age 7 (time 3) as the exposures, systolic blood pressure at age 7 as the continuous outcome, and highest parental education as the SEP-interaction term (*n* = 5,918). As the variables are added to/removed from the model they appear on the *x*-axis (with "(+)" indicating addition, and "(-)" indicating removal). The covariates/confounders constrained to be included in all models by default do not appear in this plot (SEP, maternal ethnicity, child sex and child age at outcome measurement). There appears to be little association between any of the green space hypotheses and the outcome, with model fit (deviance ratio) barely increasing as variables enter the model (see table S7 for a more formal test using relaxed and cross-validated lasso approaches). Due to collinearity with the critical period variables, the 'accumulation' hypothesis (and hence also its interaction with SEP) was dropped from this model. See figure 1 for a detailed explanation of how to interpret this plot, and table 2 for information on what each of the encoded variables mean.

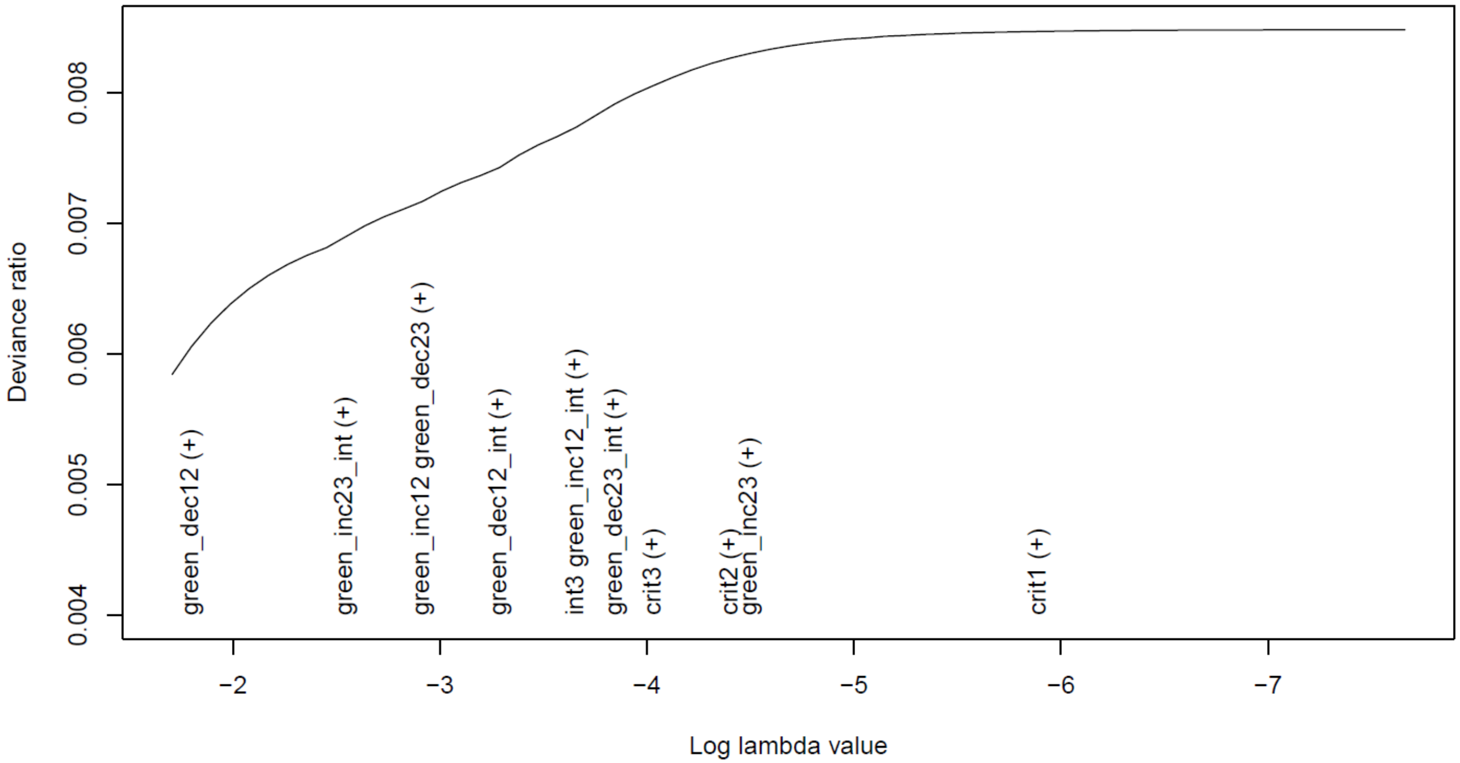

*Figure S7:* Plot of the lasso model with binary access to green space >5,000m within 300m of home in pregnancy (time 1), at age 4 (time 2) and at age 7 (time 3) as the exposures, diastolic blood pressure at age 7 as the continuous outcome, and highest parental education as the SEP-interaction term (*n* = 5,918). As the variables are added to/removed from the model they appear on the *x*-axis (with "(+)" indicating addition, and "(-)" indicating removal). The covariates/confounders constrained to be included in all models by default do not appear in this plot (SEP, maternal ethnicity, child sex and child age at outcome measurement). There appears to be little association between any of the green space hypotheses and the outcome, with model fit (deviance ratio) barely increasing as variables enter the model, although as 'green_dec12' (a decrease in green space between pregnancy and age 4) was added before all other variables, perhaps there is weak evidence that this is associated with the outcome (see table S7 for a more formal test using relaxed and cross-validated lasso approaches). Due to collinearity with the critical period variables, the 'accumulation' hypothesis (and hence also its interaction with SEP) was dropped from this model. See figure 1 for a detailed explanation of how to interpret this plot, and table 2 for information on what each of the encoded variables mean.

*References cited in supplementary information*

Boyd, A., Golding, J., Macleod, J., Lawlor, D.A., Fraser, A., Henderson, J., *et al.* (2013). Cohort profile: The "Children of the 90s"-The index offspring of the avon longitudinal study of parents and children. *Int. J. Epidemiol.*, 42, 111–127.

de Castro Pascual, M., Fossati, S., Nieuwenhuijsen, M. & Vrijheid, M. (2021). *Protocol for integrated urban environment stressors generation in LifeCycle (WP3 - Task 3.3)*.

Cole, T.J., Freeman, J. V & Preece, M.A. (1995). Body Mass Index reference curves for the UK, 1990. *Arch. Dis. Child.*, 73, 25–29.

Fraser, A., Macdonald-wallis, C., Tilling, K., Boyd, A., Golding, J., Davey smith, G., *et al.* (2013). Cohort profile: The avon longitudinal study of parents and children: ALSPAC mothers cohort. *Int. J. Epidemiol.*, 42, 97–110.

Gelman, A. (2008). Scaling regression inputs by dividing by two standard deviations. *Stat. Med.*, 27, 2865–2873.

Jaddoe, V.W.V., Felix, J.F., Andersen, A.M.N., Charles, M.A., Chatzi, L., Corpeleijn, E., *et al.* (2020). The LifeCycle Project-EU Child Cohort Network: a federated analysis infrastructure and harmonized data of more than 250,000 children and parents. *Eur. J. Epidemiol.*, 35, 709–724.

Smith, B.J., Smith, A.D.A.C. & Dunn, E.C. (2022). Statistical Modeling of Sensitive Period Effects Using the Structured Life Course Modeling Approach (SLCMA). In: *Sensitive Periods of Brain Development and Preventive Interventions* (ed. Andersen, S.L.). Springer, Cham, pp. 215–234.

Tibshirani, R.J., Taylor, J., Lockhart, R. & Tibshirani, R. (2016). Exact Post-Selection Inference for Sequential Regression Procedures. *J. Am. Stat. Assoc.*, 111, 600–620.
